# Microbiological risk ranking of foodborne pathogens and food products in scarce-data settings

**DOI:** 10.1101/2022.04.07.22273592

**Authors:** Matteo Crotta, Bhagyalakshmi Chengat Prakashbabu, Hannah Holt, Ben Swift, Paviter Kaur, Jasbir Singh Bedi, Venkata Chaitanya Pedada, Thahir Basha Shaik, Srinivasa Rao Tumati, Javier Guitian

## Abstract

In the absence of epidemiological, microbiological or outbreak data, systematic identification of the hazards and food products posing the higher risk to the consumers is challenging. It is usually in Low- and Middle-Income Countries (LMICs), where the burden of foodborne disease is highest that data tend to be particularly scarce. In this study, we propose qualitative risk-ranking methods for pathogens and food products that can be used in settings where scarcity of data on the frequency/concentration of pathogens in foodstuff is a barrier towards the use of classical risk assessment frameworks. The approach integrates the existing knowledge on foodborne pathogens, manufacturing processes and intrinsic/extrinsic properties of food products with key context-specific information regarding the supply chain(s), characteristics of the Food Business Operators (FBOs) and cultural habits to identify: (i) the pathogens that should be considered as a “High” food safety priority and (ii) the food products posing the higher risk of consumer exposure to microbiological hazards via oral (ingestion) route. When applied to the dairy sector of Andhra Pradesh (India) as a case study, *E. coli* O157:H7, *Salmonella* spp., *S. aureus* and *L. monocytogenes* were identified as a “High” food safety priority across all FBOs, *C. sakazakii* a “High” priority for the FBOs producing infant formula/milk powder whilst *Shigella* spp. and *Cryptosporidium* spp. a “High” priority when considering the FBOs operating towards the informal end of the formal-informal spectrum. The risk ranking of dairy products was informed by a preliminary cluster analysis for early identification of products that are similar with regards to intrinsic/extrinsic features known to drive the microbiological risk. Products manufactured/retailed by FBOs in the informal market were considered as posing a “High” risk for the consumers due to a widespread lack of compliance to sanitary regulations. For dairy products produced by FBOs operating in the middle and formal end of the formal-informal spectrum, the risk of consumers exposure to microbiological hazards ranged from “Medium” to “Extremely low” depending on the FBO and the intrinsic/extrinsic properties of the products. While providing risk estimates of lower resolution if compared to data-driven risk assessments, the proposed method maximises the value of the information that can be easily gathered in LMICs and provide informative outputs to support food safety decision-making in contexts where resources to be allocated for prevention of foodborne diseases are limited and the food system is complex.

## 1. INTRODUCTION

Unsafe food is responsible for a vast global burden. In 2010, the World Health Organisation (WHO) estimated that 31 foodborne biological hazards (28 microbial pathogens and 3 chemicals) were responsible for 600 million cases of foodborne illness and 33 million years of healthy life lost globally (WHO, 2015). Foodborne illnesses result from a large number of pathogen-food product combinations, making it necessary to prioritize, for purpose of surveillance and controls, those combinations that are likely to pose highest foodborne health risk (Stärk et al., 2006; Van der Fels-Klerx et al., 2018). Different frameworks have been proposed and are widely used in order to asses risk and prioritize hazards in a way that is transparent and supported by best available evidence (FAO/WHO, 2006; OIE, 2010). The risk posed by different pathogen-product combinations can be estimated quantitatively, using deterministic or probabilistic microbial risk assessment models, or qualitatively, using qualitative descriptors such as “Low”, “Medium” or “High” to describe, in non-numerical terms, the degree of belief regarding the occurrence of relevant events (e.g. whether a pathogen present in food survives a processing step) and the final risk estimate. So-called semi-quantitative approaches, in which a scoring system is used to define a logical and explicit hierarchy between the non-numerical descriptions of probability, impact, and severity, are also used for purpose of foodborne risk estimation (FAO/WHO, 2009; Van der Fels-Klerx et al., 2018). Data availability is one of the major considerations for selection of a specific approach (EFSA, 2012) with qualitative risk assessment frameworks being the usual choice when data are inadequate for quantitative assessments and expert knowledge is deemed suitable to allow differentiation between risk categories (CAC, 1999). Several examples of qualitative or semi-quantitative risk ranking of foodborne pathogens and food products are available in the literature (Van der Fels-Klerx et al., 2018). Examples range from ranking of meat-borne pathogens in intensive pork production (de Freitas Costa et al., 2020), to the ranking of chemical hazards (antibiotics) in food (van Asselt et al., 2013) or specific hazard-food combinations (Newsome et al., 2009). Recently, a risk ranking framework for food safety risks posed by emerging dietary practices has been proposed in France (Eygue et al., 2020).

Qualitative risk assessment entails a reasoned, referenced and logical discussion of the available evidence pertaining a risk, and as such, it represents a suitable framework for dealing with limited data availability. However, existing frameworks in the context of food safety rely on allocating qualitative probabilities to the frequency of the pathogen in the food or its source based on existing evidence or expert opinion. We argue that in settings such as those often encountered in Low- and Middle-Income Countries (LMICs) data on the frequency of pathogens in food are often too scarce to justify assignment of qualitative probabilities. Given that it is in LMICs where such food survey data tend to be particularly scarce or absent, that foodborne illnesses pose the highest burden, there is an urgent need for prioritization tools that do not rely on pre-existing data or knowledge on the frequency of presentation of the pathogen (Jaffee et al., 2019). Here we propose a framework to systematically and transparently assess foodborne risk in the food or its source (e.g. the animal) in the absence of data on pathogen frequency in food products. The approach, which relies on the known characteristics of the pathogen, the intrinsic and extrinsic properties of food products, their processing steps and cultural habits known to facilitate or prevent survival/growth of pathogens, also takes into consideration the socio-economic and regulatory environment within which the different Food Business Operators (FBOs) exist. While still qualitative, an assessment that is independent of pathogen frequency estimates may allow systematic prioritization in those settings where strategic resource allocation is most needed. Such an approach will avoid the need to rely on estimates of pathogen frequency in situations where they are only available from inadequate studies or from uninformed opinions and are therefore highly speculative.

The objective of this study is therefore to propose a method to qualitatively rank foodborne pathogens and food products in settings where the extremely limited/absence of data on the frequency of the pathogen prevents use of classical qualitative risk assessment frameworks. Considering that the challenge of risk prioritization in absence of pathogen frequency data is heightened for populations consuming a high variety of products, the dairy sector of Andhra Pradesh (India) is used for purpose of illustration. India is the world’s largest dairy producer where a high variety of dairy products are consumed, representing an important component of Indian culture and local diets. Within India, the state of Andhra Pradesh is the fourth largest producer of milk (NDDB, 2019).

## 2. MATERIAL AND METHODS

### 2.1. Overview of the approach

The risk assessment framework proposed in this study integrates different streams of information that can be gathered with limited resources in data-scarce settings, the general approach consists of three main steps

#### Step 1

Detailed understanding/description, achieved by means of stakeholder consultation and review of the regulatory framework, of (i) the **supply chain(s)**, (ii) the food safety **regulatory framework**, and (iii) the **risk profiles of the FBOs**.

#### Step 2

Risk ranking of **foodborne pathogens** to identify those posing the highest food safety risk. This is achieved by: (i) developing, from existing knowledge, an inclusive list of the microbiological hazards potentially posing a risk for consumers and (ii) combining this with known pathogen characteristics that shape the pathogen-specific probability of human exposure through the oral (ingestion) route from different FBOs.

#### Step 3

Risk ranking of **food products** to identify those posing the highest microbiological risk. This is achieved by: (i) describing in detail the manufacturing process of the common food products that are produced and retailed by the different FBOs, (ii) proceeding with a preliminary identification of the group(s) of products that could be considered as similar with regards to a set of variables known to drive the microbiological risk and (iii) revising this preliminary grouping by evaluating the source(s) of heterogeneity within each group.

The risk ranking of both pathogens and food products in step 2 and 3 was finalised by integrating information from step 1 to consider the specific role of the FBOs in shaping the final risk of exposure to microbial pathogens. The source and type of knowledge/information used to inform the assessment in the case study of the dairy sector of AP are outlined in figure 1.

**Figure 1.**
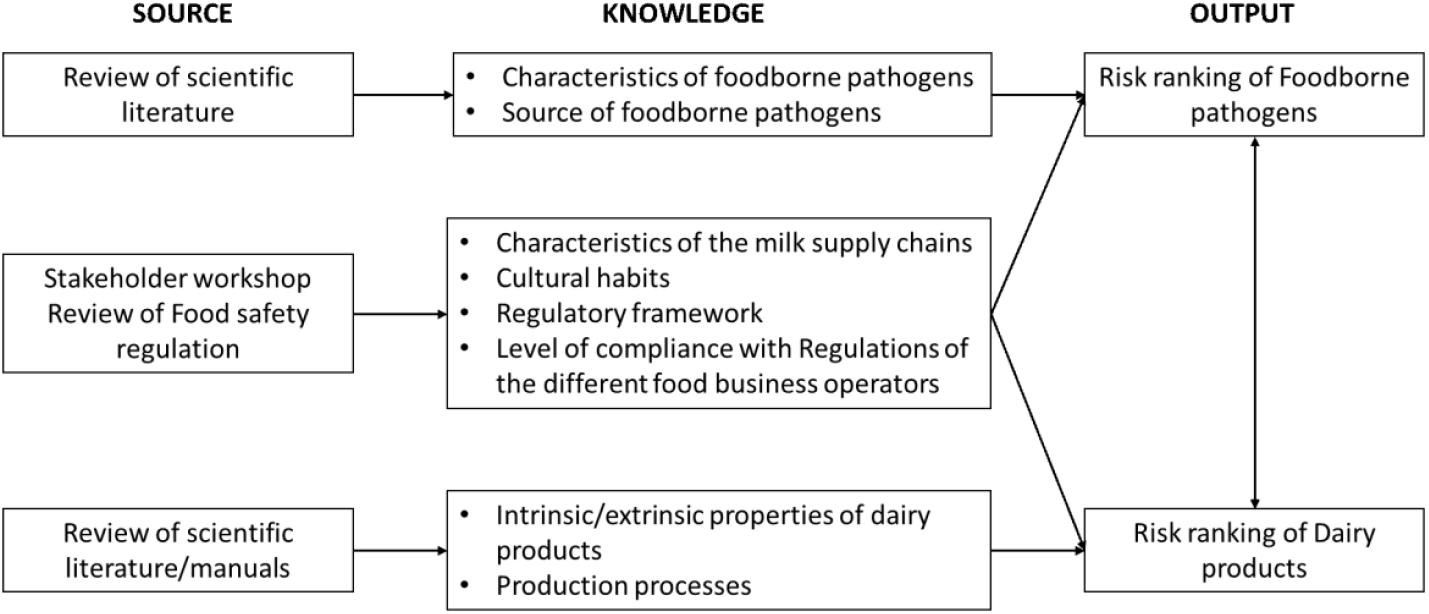
Flowchart outlining the source and the type of knowledge used to inform the qualitative risk ranking of pathogens and dairy products (output).

### 2.2. Description of the dairy supply chain, regulatory framework and food business operators

#### 2.2.1. Characteristics of the dairy supply chain in Andhra Pradesh

Most of the milk produced in Andhra Pradesh (AP) is consumed within the household with the remaining being sold through different channels involving a variety of actors operating, as in many other LMICs, at different points along a formal-informal spectrum (Blackmore et al., 2020). To understand the dairy supply chain in AP and the quantity of milk flowing through different routes and actors along the value chain, a stakeholder workshop was held with actors, or representatives of actors, at each stage in the dairy supply chain. The objectives of the workshop were to: (i) map the supply chains of the dairy sector in AP, (ii) identify the actors participating at each step of the chain and any agencies or regulations likely to influence their behaviour and (iii) gather information on key consumer habits. Full details of this exercise are provided in the Supplementary Material #1.

#### 2.2.2. Regulatory framework for dairy products in Andhra Pradesh

The key reference used to understand the food safety regulatory framework within which FBOs operate in AP was the Food Safety Standards Act, 2006 (FSSAI, 2006) and the accompanying set of Food Safety and Standards Regulations issued by the Food Safety and Standards Authority of India (FSSAI) as available on the official FSSAI website: https://fssai.gov.in/.

#### 2.2.3. Risk profile of Food Business Operators (FBOs) in the dairy chains of Andhra Pradesh

For purpose of this risk assessment, FBOs in AP are categorised according to the risks posed by lack of strict adherence to good manufacturing practices (GMP), clean-in-place (CIP) and sanitary regulations resulting in higher chances of microbial contamination of dairy products. Hence, the following ranking of FBOs (from higher risk to lower) is assumed: FBO1>FBO2>FBO3. Where FBO1 are the vendors in the informal end of the spectrum (street vendors characterised by selling or transforming and selling small volumes of milk and mobile vending arrangements); FBO2 are the small-scale manufacturers (permanent or semi-permanent small shops/kiosks) and FBO3 are the producers in the formal end of the spectrum (medium size shops, dairy companies, and cooperatives).

### 2.3. Risk profiling and ranking of foodborne pathogens

#### 2.3.1. Identification of microbiological hazards

Absence of context-specific food survey data and data on illnesses associated with pathogen-food combinations, which is the motivation of the proposed framework, precludes an *a-priori* identification of the relevant pathogens for the dairy sector of AP. Therefore, the risk ranking exercise considered pathogens known or suspected to be associated with milk and dairy products. Namely: *Aeromonas* spp., *Bacillus cereus, Brucella* spp., *Campylobacter* spp., *Clostridium botulinum, Corynebacterium* spp., *Coxiella burnetii, Cryptosporidium* spp., *Cronobacter (Enterobacter) sakazakii, Escherichia coli* O157:H7, *Leptospira, Listeria monocytogenes, Mycobacterium bovis, Salmonella* spp., *Shigella* spp., *Staphylococcus aureus, Streptococcus* spp., *Toxoplasma gondii* and *Yersinia enterocolitica*.

#### 2.3.2. Risk profiling and ranking of foodborne pathogens

The risk ranking of foodborne pathogens aimed at answering the question: *“Which foodborne pathogens represent a food safety priority in the dairy sector of Andhra Pradesh when considering specific milk and dairy products marketed by different food business operators?”*. This was addressed qualitatively by integrating knowledge regarding the biological characteristics of the pathogens (summarised in the Supplementary Material #2) with the context-specific information (Figure 1) gathered during the stakeholder workshop. For each pathogen, a risk profile summarising the key biological factors deemed relevant for the subsequent risk ranking was created using evidence from scientific literature and/or technical documents issued by public health agencies describing the hazardous properties of microbial pathogens. These included: optimal conditions for growth and survival, heat resistance, ability to produce toxins, main source of milk/dairy product contamination, infectious dose and the severity of illness. In this qualitative risk assessment, “risk” denotes the combination of the “likelihood of occurrence” and “severity of consequences” with the “likelihood of occurrence” defined as the likelihood of ingesting a dose of live bacteria deemed sufficient to result in infection via the oral (ingestion) route by consumption of dairy products. The output of the assessment is therefore a risk ranking outlining whether pathogen X should be considered as a food safety priority according to the qualitative definitions presented in the Table 1.

**Table 1.**
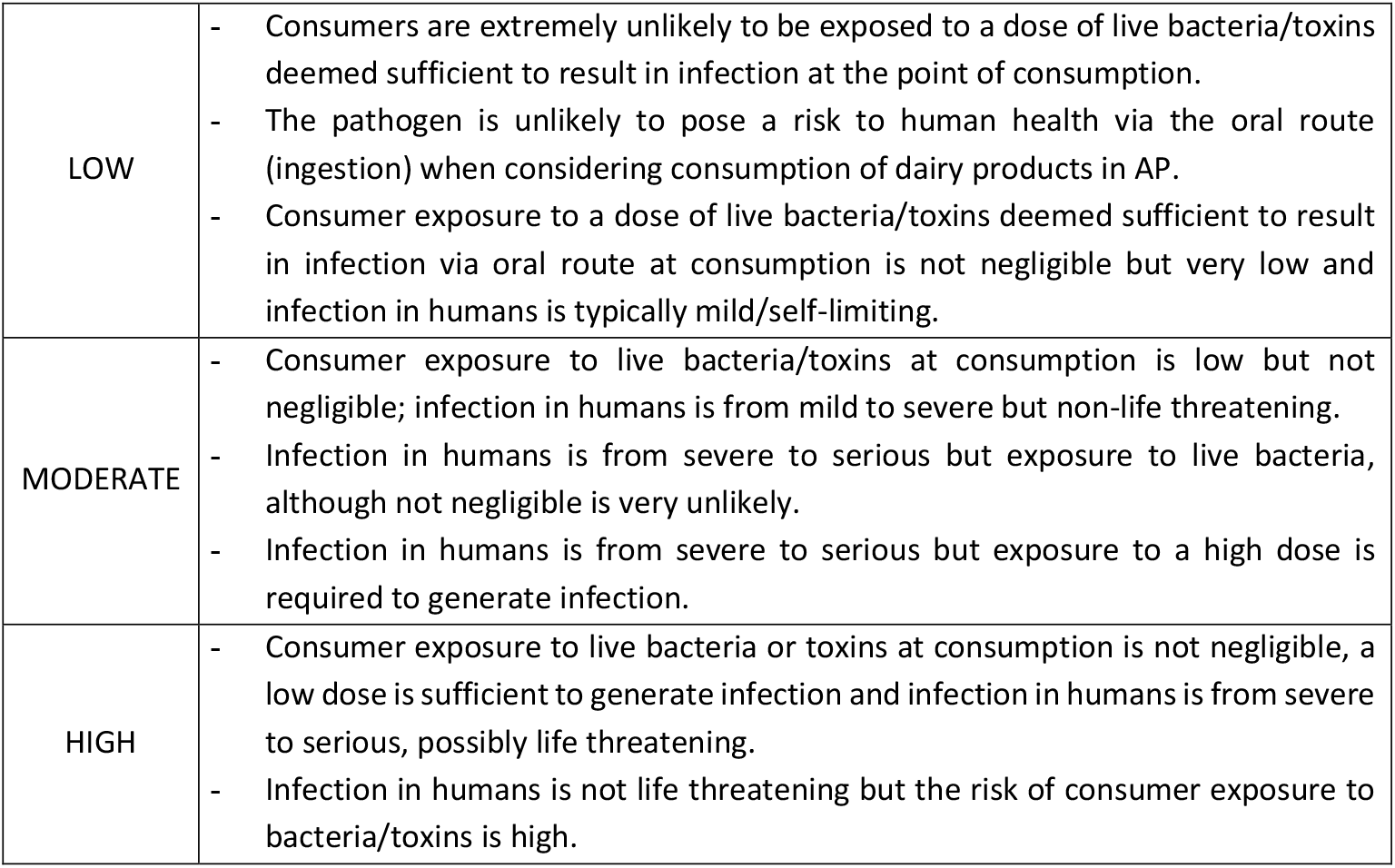
Rationale of the qualitative terms used within this risk ranking to identify each pathogen as a “Low”, “Moderate” or “High” food safety priority within the dairy sector in Andhra Pradesh.

|  |  |
| --- | --- |
| LOW | - Consumers are extremely unlikely to be exposed to a dose of live bacteria/toxins deemed sufficient to result in infection at the point of consumption.<br>- The pathogen is unlikely to pose a risk to human health via the oral route (ingestion) when considering consumption of dairy products in AP.<br>- Consumer exposure to a dose of live bacteria/toxins deemed sufficient to result in infection via oral route at consumption is not negligible but very low and infection in humans is typically mild/self-limiting. |
| MODERATE | - Consumer exposure to live bacteria/toxins at consumption is low but not negligible; infection in humans is from mild to severe but non-life threatening.<br>- Infection in humans is from severe to serious but exposure to live bacteria, although not negligible is very unlikely.<br>- Infection in humans is from severe to serious but exposure to a high dose is required to generate infection. |
| HIGH | - Consumer exposure to live bacteria or toxins at consumption is not negligible, a low dose is sufficient to generate infection and infection in humans is from severe to serious, possibly life threatening.<br>- Infection in humans is not life threatening but the risk of consumer exposure to bacteria/toxins is high. |

In the assignment of the likelihoods, full consideration was given to the contribution, in terms of expected increased risk of contamination and/or conditions favouring growth of the bacteria in food, arising from noncompliance with food safety standards by the different FBOs.

### 2.4. Risk profiling and ranking of dairy products

The risk ranking of dairy products aimed at answering the question: *“Which dairy products pose the higher microbiological risk for consumers in Andhra Pradesh when considering milk and dairy products marketed by different food business operators?”*. For purpose of this study, the most common dairy products marketed in AP were considered, namely: *UHT milk, pasteurised milk, toned milk, standardised milk, recombined milk, reconstituted milk, flavoured milk, condensed milk, khoa, basundi, burfi, peda, gulabjamun, kalajamun, milk cake, paneer, chhana-murki, rasmalai, dahi, mishti dahi, lassi, UHT lassi, yogurt, cream, ice cream, kulfi, rasgulla, junnu, kalakand, buttermilk, milk powder, junnu powder, ghee (from butter and cream)*. The assessment was performed qualitatively and was based on the intrinsic/extrinsic characteristics of dairy products and the level of compliance with food safety and sanitary regulations of the FBOs operating at different levels of the formal-informal spectrum. These aspects were considered because

i. The products are very different from each other but can be characterised by well-established intrinsic and extrinsic factors known to favour/prevent bacterial contamination and growth/survival within a food matrix.
ii. Many dairy products are made and can be purchased from different FBOs, such as street vendors, kiosks, small shops or supermarkets. While the manufacturing processes follows the same steps and the biochemical characteristics of the dairy products can assumed to be comparable, the same product made by different FBOs may pose very different risk of microbiological contamination arising from the environment, unhygienic handling or retail form (i.e. loose form, manually or industrially packaged).

Intrinsic factors are the inherent (natural or artificially occurring) physical, chemical or biological characteristics of the food matrix such as water activity (aw), pH, availability of nutrients or antimicrobial components while extrinsic factors are those controlled by the external processing conditions such as thermal treatments, manipulation or preservation methods (Demirci et al., 2020). In this particular assessment (dairy products in AP), given the high variety of products commonly available to consumers, characterisation of dairy products was initially informed by hierarchical cluster analysis aimed at identifying groups of dairy products that can be considered as similar in terms of intrinsic and extrinsic characteristics driving the microbiological risk.

The final ranking was then finalised considering the additional risk of contamination arising from the products being manufactured by the different FBOs.

#### 2.4.1. Characterisation of dairy products

The manufacturing process of each dairy product was first reviewed and summarised into a manufacturing table (Supplementary Material #3); then, for each product, the information related to key intrinsic and extrinsic properties known to favour or prevent microbial presence/growth were described according to the variables

(i) Initial Heat treatment (IHT), (ii) Water activity (aw), (iii) pH & Starter culture and (iv) Final Heat Treatment (FHT). Detailed justification of why these variables were selected and how they were measured is provided in Supplementary Material #1.

#### 2.4.2. Cluster analysis

Hierarchical Clustering on Principal Components (HCPC) was used to construct a hierarchical tree showing links between dairy products or groups of dairy products based on the variables mentioned in 2.4.1. As all variables were categorical, hierarchical classification of dairy products was based on the principal components obtained by means of Multiple Correspondence Analysis (MCA) (Greenacre & Blasius, 2006). Briefly, MCA provides a graphical representation of the data by creating synthetic independent dimensions to describe the relationships between the levels of the variables used to describe the objects (i.e. the products). The dairy products are therefore projected onto these dimensions at a distance where the variability of the projected points (projected inertia) is maximised. As a result, two products will be shown close to each other if they share a relatively large number of characteristics or far apart if they have very different profiles.

The MCA was initiated keeping all the dimensions and the ideal partitioning of the hierarchical tree determining the final number of clusters of dairy products was done selecting the number of clusters “*n”* for which the loss of inertia is minimal when passing from “*n”* to “*n+1”*. Results of the cluster analysis provided a visual representation (dendrogram) of the similarities/dissimilarities between groups of dairy products in terms of characteristics favouring or preventing microbial growth/survival. Analyses were done using the “FactoMineR” package (Lê et al., 2008) in R software.

#### 2.4.3. Risk ranking of dairy products

Objective of the cluster analysis described in 2.4.2. was to group products into relatively homogeneous groups based on a set of variables. However, as some dissimilarities may still exist within the products in each cluster; from the results of the cluster analysis the final risk ranking was finalised evaluating the food safety impact of the features determining dissimilarities amongst the products within each group (if any). In addition, for each product, the risk of microbiological contamination arising from noncompliance to food safety Regulation and hygienic standards was also considered by assuming that if the same product is manufactured/retailed by different FBOs (i.e. FBO1, FBO2 and FBO3 as specified in 2.2.3) the higher risk of consumer exposure to microbiological hazards is posed by products purchased from FBO1 followed to FBO2 and then FBO3.

The output of the risk assessment for dairy products is therefore a risk ranking outlining whether product X should be considered as a dairy product that for its intrinsic/extrinsic characteristics in combination with the risk profile of the FBO poses a risk of exposure to foodborne pathogens according to the qualitative definitions presented in the Table 2.

**Table 2.** Rationale of the qualitative terms used within this risk ranking to identify each dairy product as posing an “Extremely low”, “Very Low”, “Low”, “Moderate” or “High” risk for consumers of dairy products in Andhra Pradesh.

|  |  |
| --- | --- |
| EXTREMELY LOW | Regardless of whether the intrinsic/extrinsic characteristic of the product are favourable for microbial growth, there is a final heat treatment deemed sufficient to eliminate foodborne pathogens before hermetic packaging; strict adherence to food safety regulations, GMP, CIP are systematically in place to minimise chances of environmental contamination. |
| VERY LOW | Regardless of whether the intrinsic/extrinsic characteristic of the product are favourable for microbial growth, there is an initial treatment deemed sufficient to eliminate foodborne pathogens present in milk but not a final heat treatment. However, the manufacturing process is highly standardised; strict adherence to food safety regulations, GMP, CIP to minimise chances of environmental contamination. |
| LOW | The intrinsic/extrinsic characteristic of the product are not favourable for microbial growth; although there is an initial or final heat treatment deemed sufficient to eliminate foodborne pathogens, the product is sold in loose form or manually packaged. Food safety standards are not always met and GMP/CIP not systematically adopted by the FBO; environmental/human cross-contamination is possible but growth is prevented. |
| MODERATE | The intrinsic/extrinsic characteristic of the product are favourable for microbial growth; although there is an initial or final heat treatment deemed sufficient to eliminate foodborne pathogens, the product is sold in loose form or manually packaged. Food safety standards are not always met and GMP/CIP not systematically adopted by the FBO; environmental/human cross-contamination is possible and growth is not prevented. |
| HIGH | There is an initial and/or final heat treatment deemed sufficient to eliminate foodborne pathogens but the product is often home-made, sold in loose form or manually packaged. Food safety standards are rarely met, GMP/CIP not adopted by the FBO and microbiological contamination of the product is very likely. The risk |
|  | should be considered high regardless of whether the intrinsic/extrinsic characteristic of the product are favourable for microbial growth. |

## 3. RESULTS

### 3.1. Milk supply chains in Andhra Pradesh, food business operators and regulatory framework

During the mapping exercise, a description of the milk flows from production to consumption was built with all participants using a consensus-based approach. The resulting Sankey diagram (Figure 2) demonstrates the complexity of the system with a large number of actors involved.

**Figure 2.**
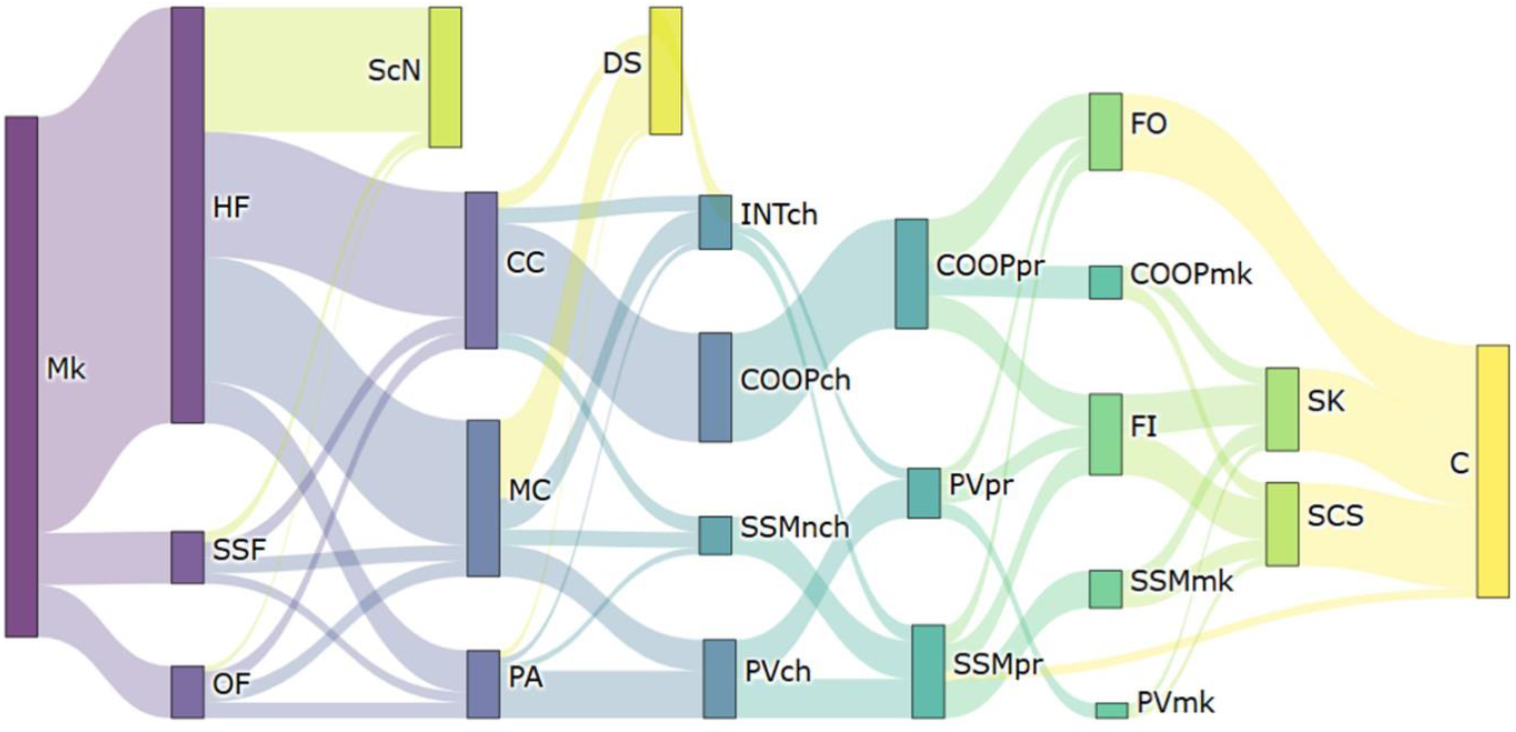
Sankey diagram outlining the flow of milk in Andhra Pradesh from production to consumption quantitatively. The width of the arrows connecting the different nodes is proportional to the total amount of milk flowing along that route. Mk=Milk, HF=Household farm, SSF=Small-scale farm, OF=Organised farm, ScN= Self-consumption and neighbourhood, MC=Milk collector, CC=Collection centre, PA=Private agent, DS=Direct sale, INTch=Intermediary (chilling), COOPch (Cooperatives (chilling), PVch=Private dairy company (chilling), SSMnch=Small scale manufacturers (NOT chilled), COOPpr=Cooperatives (processing), SSMpr=Small scale manufacturer (processing), PVpr=Private dairy company (processing), FI=Food industry, FO=Food business operators, COOPmk=marketed by cooperatives, SSMmk=marketed by small scale manufacturer, PVmk=marketed by private dairy companies, SK=Supermarket, SCS=Street corner shops, C=Consumers.

A full description of the milk supply chains in Andhra Pradesh is presented in Supplementary Material #1. The tool used to visually summarize the flow of milk quantitatively has been made available as a web-based application (https://mcrvc.shinyapps.io/riverflows/).

### 3.2. Risk ranking of foodborne pathogens

Results of the final risk ranking of foodborne pathogens and brief rationale are presented in Table 3. The full risk profile describing each pathogens’ characteristics that was used to rank the pathogens is presented in the Supplementary Material #2.

**Table 3.** Risk ranking of milk-borne pathogens that are judged to be a Low (L), Moderate (M) or High (H) food safety priority in the dairy sector in Andhra Pradesh when considering the oral (ingestion) route of infection and dairy products supplied by different types of Food Business Operators (FBO1, FBO2, FBO3) and rationale.

| Pathogen | Rationale | FBO1 | FBO2 | FBO3 |
| --- | --- | --- | --- | --- |
| *Aeromonas* spp. | The pathogen is not shed in milk, does not survive pasteurisation and infection is typically self-limiting. The infectious dose is high ($>10^9$ CFU) and the foodborne route is deemed to have marginal contribution to the global burden of disease, compared to exposure via contaminated water. Dairy products do not seem to pose a high risk for the presence of the bacteria compared to other foodstuffs. | L | L | L |
| *Brucella* spp. | Infection in humans is severe and prevalence of infection in cattle in India is non-negligible, the pathogen does not survive pasteurisation but can survive for long time in milk and fermented dairy products. While certainly a priority if considering occupational exposure, consumption of raw milk and products made with raw milk is negligible in Andhra Pradesh, hence, infection via the oral route (ingestion) is in principle unlikely. The risk of human exposure to *Brucella* spp. cannot however be entirely excluded in case of domestic heat-treatments resulting in decimal reductions not sufficient to eliminate the bacteria from the raw milk before consumption should the pathogen be present in high amounts. Thanks to the cultural habits and conditions needed to result in exposure of live bacteria (i.e. high level of contamination that makes the heat-treatment ineffective), chances for infection via oral route are likely to be very low. However, infection is severe and for this reason the risk of *Brucella* spp. infection via oral route is considered "moderate" for milk and dairy products sold by actors in the informal end of the spectrum. | M | L | L |
| *Campylobacter* spp. | The pathogen is part of the intestinal flora of various farm and companion animals and can be present in milk because of faecal contamination or cross contamination with meat of other animals, particularly poultry. *Campylobacter* infections are rarely life threatening; however, the infectious dose is relatively low. The pathogen is very heat-sensitive and does not survive pasteurisation. Considering consumption of raw milk and products made with raw milk is negligible in AP; infection via the oral route (ingestion) is highly unlikely in settings where the risk of post-processing cross-contamination from other food sources or surrounding environment is minimised. | M | L | L |
| *Corynebacterium* spp. | Infections from zoonotic strains of *Corynebacterium* are not primarily associated with handling of infected dairy products/companion animals or consumption of contaminated raw milk. The pathogen does not survive pasteurisation. | L | L | L |
| *C. botulinum* | The pathogen is ubiquitous and has been detected in a variety of food including milk. While infection can be life threatening, this is dependent upon ingestion of pre-formed neurotoxins. Conditions for toxin production are very specific (i.e. temperature abuse of non-acidic anaerobic environment in absence of competitive bacteria) and not met in dairy products commonly produced in AP. | L | L | L |
| *C. burnetii* | The pathogen is shed in milk of infected cows as an obligate intracellular microorganism, it does not grow in milk and is eliminated by pasteurisation. The main route of infection seems to be airborne and evidence of the pathogen as a milk-borne hazard capable of causing disease through via oral route (ingestion) is weak. If any, the main risk is represented by consumption of highly contaminated raw milk which is negligible in AP. | L | L | L |
| *Leptospira* | Shed in urine of infected animals, leptospirosis is considered an occupational disease for which the main route of infection is percutaneous through contact with contaminated water or damp soil. While contamination of milk cannot be entirely excluded it is restricted to cross-contamination with infected urine. Milk and dairy products are not a suitable medium for growth of this pathogen, if any, the main risk is represented by consumption of highly contaminated raw milk which is negligible in AP. | L | L | L |
| *M. bovis* | While bovine tuberculosis in humans is uncommon in Europe and other high-income countries, it is still a major public health concern in India. As for *Brucella* spp., bovine tuberculosis remains a public health priority within the dairy sector in India and AP. Although the pathogen can be shed in the milk of infected animals, it does not survive pasteurisation and considering consumption of raw milk and products made with raw milk is extremely uncommon in AP, infection via oral route (ingestion) is highly unlikely. Existing evidence suggest that infection via the oral route requires thousands or millions of organisms as compared to less than 10 through inhalation, however, as for *Brucella* spp., because of the cultural habits and conditions needed to result in exposure of live bacteria (i.e. high level of contamination that makes the heat-treatment ineffective), chances for infection via oral route are likely to be very low. However, infection can result in severe illness and for these reasons, the risk of *M. bovis* infection via oral route is considered as “moderate” for milk and dairy products sold by actors in the informal end of the informality spectrum. | M | L | L |
| *Streptococcus* spp. | Most streptococcal illnesses in people are caused by species normally maintained in humans (e.g. *S. pneumoniae*, *S. pyogenes* or human-adapted strains of *S. agalactiae*) and originate from opportunistic infection in the host or transmission resulting from close physical contacts. Ingestion of undercooked pork (e.g., *S. suis*), horsemeat (*S. equi* subsp. *Zooepidemicus*), fish (*S. agalactiae* ST283) and unpasteurised dairy products (*S. equi* subsp. *Zooepidemicus*), have however been associated with infection in humans. The genetic profile of *S. agalactiae*, the major cause of bovine mastitis, seems to be distinct from the strain causing infection in humans; likewise, there is no evidence of milk-borne human infections due to *S. uberis* or *S. dysgalactiae* subsp. *Dysgalactiae*, other strains responsible for mastitis in cattle. With the advent of pasteurisation, the incidence of streptococcal outbreaks has drastically reduced; recent outbreaks have mainly involved *S. pyogenes* and food products different from milk or dairy products. | L | L | L |
| *T. gondii* | While consumption of raw goats' milk is often identified as a risk factor for human toxoplasmosis, the risk of acquiring infection by drinking cow's milk, if any, seems to be minimal. As an obligate intracellular coccidian, *T. gondii* does not replicate in food, as such, the risk pathway for milk-borne toxoplasmosis requires consumption of raw milk, which is negligible in AP. | L | L | L |
| *Y. enterocolitica* | The pathogen is ubiquitous and can grow on a variety of foods, even at refrigeration temperatures, however, it is killed by pasteurisation. Infection is not life threatening and infectious dose is relatively high ($10^6$-$10^8$). The primary source of human yersiniosis is pigs and while *Y. enterocolitica* is occasionally detected in heat-treated milk products, contamination is mainly environmental. Dairy products do not seem to pose a high risk for the presence of the bacteria compared to other foods such as pork or beef. | L | L | L |
| *B. cereus* | Although this pathogen is frequently detected in various foods including milk, most *B. cereus* outbreaks have involved food commodities other than milk and dairy products. This is probably because conditions for *B. cereus* to generate infection are unlikely to be met in dairy products before spoilage. The risk seems to be limited to pasteurised dairy products where the microorganism can multiply (or spores are already present) at high concentration ($>10^5$ CFU/g) without spoiling the product such as extended shelf-life chilled products, desserts or reconstituted powdered milk. | M | M | M |
| *Shigella* spp. | The bacteria does not survive pasteurisation and infection from consumption of dairy products is likely to be sporadic, mainly due to poor personal hygiene and handling of processed dairy products. However, the infectious dose is believed to be very low (as few as 10 CFU) therefore shigellosis is a very easily transmitted disease. Clinical signs include diarrhoea that can last from a few days to several weeks. As a pathogen for which the only known reservoirs are humans and large primates, the main transmission route is via food contaminated with human faeces. Considering GMP and CIP are not systematically adopted by processors in the informal sector, *Shigella* spp. should be considered a high food safety priority in this setting. | H | H | L |
| *C. sakazakii* | While the main reservoir for *C. sakazakii* remains unknown, the pathogen is ubiquitous in food and processing environments. Its ability to form biofilms promoting adherence and environmental stress resistance together with the peculiar ability of surviving on dry substrates for a long time make this pathogen a serious threat for the dairy industry; particularly milk powder, powder infant formula and milk protein-producing facilities. | L | L | H* |
| *Cryptosporidium* spp. | *Cryptosporidium* spp. is considered the main waterborne parasite worldwide due to its ability to survive wastewater treatment and drinking water disinfectants. The parasite is excreted with faeces of infected animals, the main risk for human exposure in the dairy sector is via consumption of raw milk and dairy products made with raw milk but also, and more relevant for AP in settings where CIP is not rigorously applied, is the use of utensils and equipment cleaned with contaminated water. | H | H | L |
| *E. coli* O157:H7 | The Shiga toxin-producing serotype *E. coli* O157:H7 is part of the intestinal flora of ruminants and presence in milk is due to faecal contamination. Infectious dose is low, ranging from 10 to 100 CFU and infection in humans can be severe and life threatening. Key risky dairy products are raw milk and in general, any processed dairy product with a risk of post-pasteurisation faecal contamination via human handling. | H | H | H |
| *L. monocytogenes* | This ubiquitous pathogen can contaminate milk and dairy products at several stages of the food chain via environmental cross-contamination. *L. monocytogenes* has been isolated in a variety of dairy products, it does not survive pasteurisation but can grow at refrigeration temperature and adapt to acidic environments. Infection in humans can be life threatening and milk and dairy products are frequently implicated in listeriosis outbreaks. | H | H | H |
| *Salmonella* spp. | *Salmonella* spp. is part of the intestinal flora of ruminants and presence in milk is typically due to faecal contamination. Key risky dairy products are raw milk and in general, any processed dairy product with a risk of post-pasteurisation faecal contamination through human handling. Infectious dose seems to be rather high ($10^5$ CFU) but *Salmonella* infections in humans are severe although rarely life threatening. | H | H | H |
| *S. aureus* | *S. aureus* is one of the major causes of bovine clinical and sub-clinical mastitis. Infection in humans is moderate and self-limiting resulting from ingestion of pre-formed and thermo-stable enterotoxins. Methods for detection of subclinical mastitis in milking animals are not routinely used in India and Andhra Pradesh, and the risk of *S. aureus* being present in milk at high amounts before heat treatment is not negligible. Humans are also known reservoirs and post-pasteurisation contamination via handling is not uncommon. | H | H | H |

The pathogens identified as a “High” food safety priority across all FBOs were: *E. coli* O157:H7, *Salmonella* spp., *S. aureus* and *L. monocytogenes* whilst *B. cereus* was identified as a “Moderate” food safety priority. *C. sakazakii* was identified as a “High” food safety priority for FBO3, but this is limited to the industrial production of infant formula/milk powder (not produced by FBO1 and FBO2). On the other hand, *Shigella* spp. and *Cryptosporidium* spp. were identified as a “High” food safety priority for FBO1 and FBO2; this is because the risk arising from these pathogens is mainly related to unhygienic handling of food and adoption of GMP and CIP principles by these FBOs is very limited. *M. bovis* and *Brucella* spp. are endemic in cattle in India and AP, therefore herd prevalence is non-negligible, however, the oral (ingestion) route was considered in principle as unlikely to be a relevant exposure pathway for these pathogens. This is because effective pasteurisation is deemed sufficient to eliminate the risk, should these pathogens be already present in the milk of infected animals. However, it was also considered that domestic heat-treatment of raw milk directly purchased from the neighbouring farm, milkman or collection centre might not be sufficient to achieve a log reduction such as to completely eliminate the pathogens should these be present at high concentrations. Although chances of exposure to live bacteria via oral route are very low, considering the severity of the diseases, *M. bovis* and *Brucella* spp. were considered of “Moderate” priority.

### 3.3. Risk ranking of dairy products

#### 3.3.1. Cluster analysis

Altogether, the first four dimensions of the MCA explained 93.1% of the variance; based on inertia criterion the increase in between-cluster inertia when moving from 5 to 6 clusters (figure 3) was minimal, compared to from 4 to 5, it was hence decided to partition the hierarchical tree in 4 clusters.

**Figure 3.**
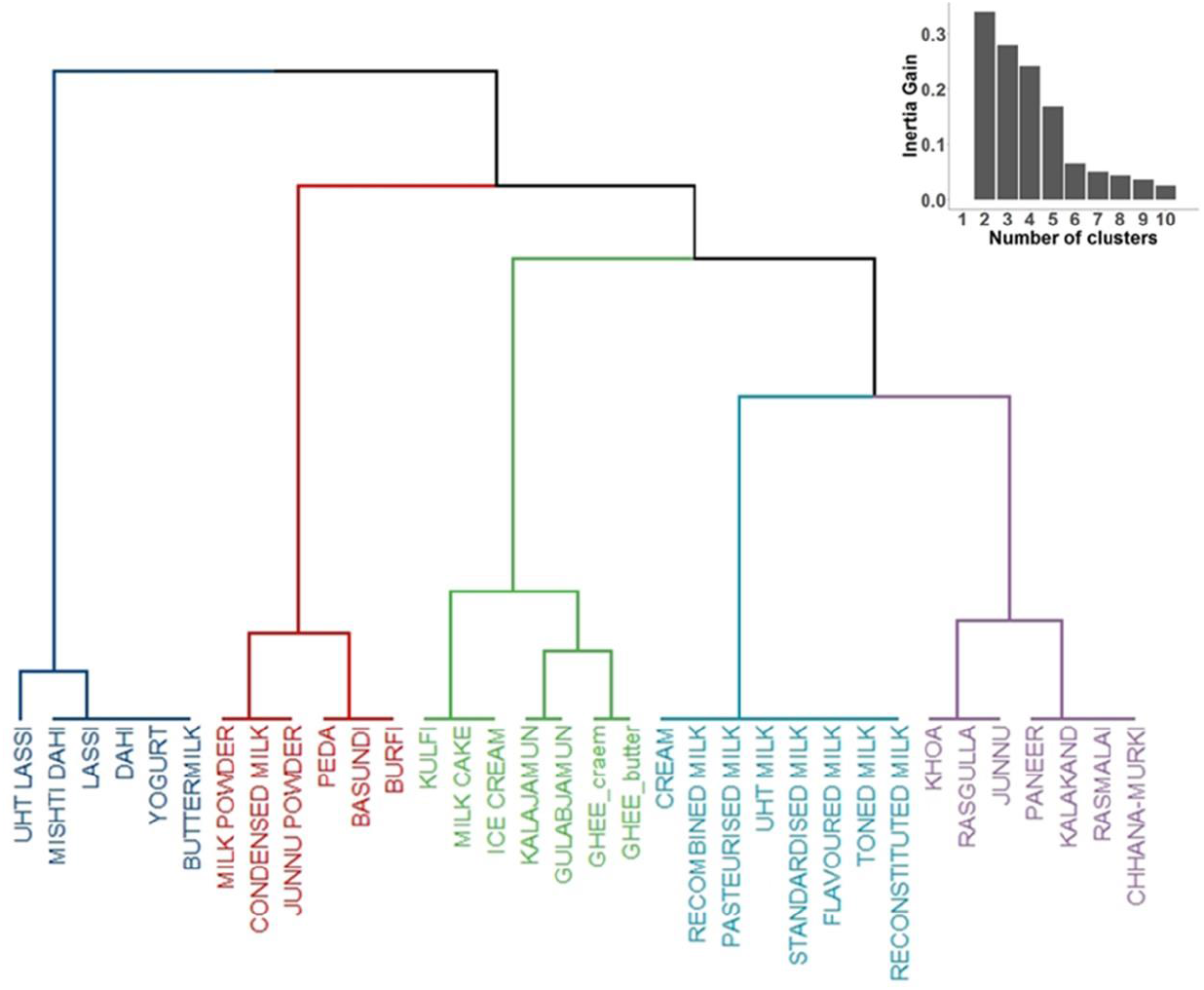
dendogram showing the hirarchical clustering of dairy products and the barchart of the amount of inertia gained when moving from n clusters to n+1.

Products in the first cluster: UHT lassi, mishit dahi, lassi, dahi, yogurt and buttermilk are all fermented products of high aw that are not heat-treated before packaging (exception made for UHT lassi). In fact, all these products are characterised by the addition of a starter culture after an initial heat treatment, the risk arising from the possible microbial contamination is mitigated by presence of Lactic Acid Bacteria (LABs) that proliferate during fermentation creating an unfavourable and competitive environment for most of the pathogenic bacteria.

Products in the second cluster: powder milk/junnu, condensed milk, peda, burfi and basundi all have low aw and undergo an initial heat treatment as part of the manufacturing process. While powdered products and the condensed milk also undergo a final heat treatment before packaging this is not the case for the Indian sweets basundi, burfi and peda.

The third cluster was the most heterogeneous and included: kulfi, milk cake, ice cream, gulabjamun, kalajamun and ghee (butter and cream methods). All these products are characterised by medium aw values suboptimal for growth of some pathogens such *Shigella, E. coli* O157:H7 or *Salmonella* spp. but still optimal for other such as. *L. monocytogenes* or *S. aureus*. Gulabjamun and kalajamun are khoa-based sweets not undergoing any initial heat treatment but are heat treated at T>100°C before packaging. Similarly, ghee is made from either pasteurised cream or butter treated since the beginning at T between 100 and 110°C before packaging.

Products in the fourth cluster included: cream, reconstituted milk, pasteurised milk, UHT milk, standardised milk, flavoured milk, toned milk and recombined milk. Products in this group are manufactured through a processing circuit that is completely closed with minimal risk of cross-contamination from the environment, have high aw but are at least pasteurised before packaging.

Finally, products in the fifth cluster: khoa, rasgulla, junnu, paneer, kalakand, Rasmalai and channa-murki are characterised by having an initial heat-treatment as part of the processing and high aw. Junnu, rasgulla and khoa are subjected to a prolonged heat treatment at high temperatures before packaging.

#### 3.3.2. Risk ranking of dairy products

The risk ranking of dairy products was performed integrating: (i) the results of the clustering, (ii) the characteristics of the individual products resulting in within-cluster dissimilarities where needed, and (iii) the risk profile of the FBOs where products can be purchased. Results of the final risk ranking are summarised in Table 4.

**Table 4.** Risk ranking of dairy products. Dairy products are classified as “Extremely low”, “Very low”, “Low”, “Moderate” or “High” risk of consumer exposure to microbiological hazard; classification is informed by integrating the intrinsic/extrinsic characteristics of the products with the additional risk of microbiological contamination arising from the FBO. n.a.=product not normally produced/retailed by the FBO

| PRODUCT | FBO1 | FBO2 | FBO3 |
| --- | --- | --- | --- |
| Buttermilk | HIGH | MEDIUM | VERY LOW |
| Chhana-murki | HIGH | MEDIUM | VERY LOW |
| Kalakand | HIGH | MEDIUM | VERY LOW |
| Paneer | HIGH | MEDIUM | VERY LOW |
| Flavoured milk | HIGH | MEDIUM | EXTREMELY LOW |
| Junnu | HIGH | MEDIUM | EXTREMELY LOW |
| Khoa | HIGH | MEDIUM | EXTREMELY LOW |
| Burfi | HIGH | LOW | VERY LOW |
| Dahi | HIGH | LOW | VERY LOW |
| Ice cream | HIGH | LOW | VERY LOW |
| Kulfi | HIGH | LOW | VERY LOW |
| Lassi | HIGH | LOW | VERY LOW |
| Mishti dahi | HIGH | LOW | VERY LOW |
| Peda | HIGH | LOW | VERY LOW |
| Gulabjamun | HIGH | LOW | EXTREMELY LOW |
| Ghee (butter) | HIGH | n.a. | n.a. |
| **Milk cake** | n.a. | MEDIUM | VERY LOW |
| **Rasmalai** | n.a. | MEDIUM | VERY LOW |
| **Yogurt** | n.a. | MEDIUM | VERY LOW |
| **Cream** | n.a. | MEDIUM | EXTREMELY LOW |
| **Rasgulla** | n.a. | MEDIUM | EXTREMELY LOW |
| **Junnu powder** | n.a. | LOW | EXTREMELY LOW |
| **Kalajamun** | n.a. | LOW | EXTREMELY LOW |
| **Milk powder** | n.a. | LOW | EXTREMELY LOW |
| **Ghee (cream)** | n.a. | LOW | EXTREMELY LOW |
| **Basundi** | n.a. | LOW | VERY LOW |
| **Condensed milk** | n.a. | n.a. | EXTREMELY LOW |
| **Pasteurised milk** | n.a. | n.a. | EXTREMELY LOW |
| **Recombined milk** | n.a. | n.a. | EXTREMELY LOW |
| **Reconstituted milk** | n.a. | n.a. | EXTREMELY LOW |
| **Standardised milk** | n.a. | n.a. | EXTREMELY LOW |
| **Toned milk** | n.a. | n.a. | EXTREMELY LOW |
| **UHT lassi** | n.a. | n.a. | EXTREMELY LOW |
| **UHT milk** | n.a. | n.a. | EXTREMELY LOW |

From definitions given in Table 2, in principle, all products manufactured by FBO1 represent an “extremely low” to “very low” risk of exposure to the microbiological hazards for the consumers. However, when the same products are manufactured/distributed by FBO2 the risk increases to “low” or “medium”; chances for microbial contamination are assumed to be higher for products manufactured by FBO2 and the difference between “low” and “moderate” is mainly linked to the intrinsic characteristics of the product favouring microbial growth.

A further increase in the risk is assumed for the same products manufactured by FBO3, where chances for environmental/human contamination drive the risk estimate in such a way that the intrinsic characteristics favouring/preventing microbial growth become irrelevant.

## 4. DISCUSSION

The intent of this study was to propose a method for systematic risk ranking of foodborne pathogens and food products in settings where data on the frequency/concentration of pathogens in foods are largely absent. In these settings, even the assignment of qualitative probabilities to the minimum set of events along the risk pathways until consumption (i.e. probability of the food product being contaminated or the probability of the pathogen surviving processing) would be highly speculative and unjustified.

The main advantage of the approach presented here is that it is entirely based on existing knowledge and information that can be easily gathered in any setting. However, a limitation is that the prioritization may be obtained at a low level of resolution leading to the identification of a *group* of pathogens or *group* of products posing the highest risk for the consumers. The choice of the probability scales from “Low” to “High” for pathogens and “Extremely low” to “High” for products were a compromise between a desired practically informative level or resolution of the final outputs and the level of discrimination that could be realistically achieved from the available information. However, if supported by rigorous, comprehensive and logical reasoning, this qualitative prioritization of groups of pathogens and products can be highly informative to support decision-making; particularly if resources to be allocated for the prevention of foodborne diseases are limited, the food system is complex and food-safety decisions need to be made. The potential of this risk ranking approach has been demonstrated by applying it to the complex dairy sector of AP, where milk flows through a network of formal and informal actors resulting in very diverse dairy products being available to consumers. Risk ranking of pathogens was performed through a logical discussion of the exposure pathways as opposed to survey data. This is because (i) there are very limited data on the presence or absence of pathogens in milk and dairy products in India in general and AP specifically, (ii) results from other parts of India/countries are not necessarily representative of AP and (iii) some potentially relevant pathogens are not targeted by microbiological surveys.

When considering the case study of the AP dairy sector, the risk ranking of pathogens identified *E. coli* O157:H7, *Salmonella* spp., *S. aureus* and *L. monocytogenes* as the pathogens that should be regarded as high food safety priority and *B. cereus* as a moderate priority for all FBOs. *Shigella* spp. was identified as high priority for FBO1 because the risk is restricted to cross-contamination with human faeces and hence unhygienic handling or production in highly unregulated settings but the infectious dose is extremely low (Zaidi & Estrada-García, 2014). Similarly, *Campylobacter* spp. and *Cryptosporidium* were considered as moderate food safety priority for FBO1 and FBO2 due to the risk mainly arising from unhygienic handling of food and very limited adoption of sanitary measures by these FBOs. On the other hand, *C. sakazakii* was identified as a high food safety priority for FBO3 due to the risk posed by this pathogen when present in milk powder and powdered infant formula (not produced by FBO1 and FBO2). The fact that important zoonotic diseases such as bovine tuberculosis and brucellosis are still prevalent in the country (Mangtani et al., 2020; Refaya et al., 2020) might initially cause alarm in terms of consumer exposure to these important zoonotic pathogens. Our assessment based on the integration of different streams of information, has identified these pathogens as of “moderate” priority but only for a specific segment of the population (i.e. those purchasing raw milk for self-consumption directly from neighbouring farms, milkman or collection centre) and under a specific set of circumstances (i.e. inadequate domestic treatment of highly contaminated raw milk). Consumers in AP reportedly always boil milk before consumption, and dairy products are made from pasteurised (or equivalently heat-treated milk) in both the formal and informal supply chain. These are key behaviours, which minimise the risk arising from pathogens that would be considered as a high food safety priority otherwise.

Context-specific qualitative information played a major role in the risk ranking of foodborne pathogens and repeating this exercise in other settings characterised by different consumer’ habits (Chengat Prakashbabu et al., 2020), will probably result in a very different set of priority pathogens. Of note that the method proposed here should not be considered as relevant only for LMICs. In fact, the same approach can be used for early identification of potential hazards at which consumers (or group of consumers) can be exposed to because of specific habits, believes or changes in consumption trends (FSA, 2018; Golden et al., 2022; Tomasevic et al., 2018). For this reason, integration of different streams of information as proposed in this study is essential to minimise biases in the qualitative characterisation of the risk.

The risk ranking of food products was based on methods for classification and evaluation of similarities/dissimilarities between products or group of products based on the intrinsic and extrinsic factors known to favour or prevent microbial contamination and/or growth integrated with the expected risk of microbiological contamination that characterises the different FBOs. Consideration of the two elements is particularly relevant in settings such as AP or any other LMICs where very diverse FBOs coexist and the same products might pose a very different food safety risk depending of the manufacturer/distributor.

The concise nature of the preliminary cluster analysis inevitably required a further within-cluster evaluation of dissimilarities between products within and across clusters for the scope of the final risk ranking. However, the visual representation of the hierarchies together with the inspection of features characterising each cluster facilitates identification of groups of products that could be quickly considered of low (or high) risk. This preliminary step can in fact be very useful if a long list of food products is to be evaluated, as is the case in the Indian dairy sector. If epidemiological data are available, other evidence-based approaches can certainly be used (Sumner et al., 2005; Xavier et al., 2014); the method proposed here finds its value precisely in contexts where data are scarce/absent and a high variety of food products are marketed. In such case, a risk ranking simply based on few but well-established intrinsic and extrinsic properties can serve to narrow down the spectrum of food products and consequently, support a more efficient allocation of resources towards those that are more likely to pose the higher risk for the consumers.

The factors included in the analysis were those relevant for the common dairy products marketed in AP. Should this exercise be repeated for other food commodities where manufacturing processes includes other important intrinsic or extrinsic factors (e.g. ageing, inclusion of chemical preservatives as part of the product formulation or packaging under modified atmosphere), these should of course be considered for either the classification by means of cluster analysis and the final ranking.

It should be noted that the final ranking of food products was intentionally made pathogen non-specific as this was judged impractical. Combining the ranking of pathogens with the ranking of products would in fact require a higher level of resolution of the output allowing to distinguish for example the risk arising from a product judged to pose a “high” risk for pathogens considered as “high” food safety priority and pathogens considered as “moderate” or “low” priority. Considering the limited set of information on which this framework is based, attempting such specific product-pathogen ranking would have led to an output surrounded by high uncertainty. Considering the main objective of the proposed methods is to inform decision-making in data-scarce settings, the most practical option is to accept the compromise of an output that is of low resolution but informative. Hence, the more practical option for an early pathogen-product interpretation of the risk is to consider the ranking of products to identify the FBO-product combinations that should be given priority if food safety risk-based monitoring/surveillance plans are to be implemented.

Qualitative risk assessments based on logical appraisal of the available evidence provide conclusions that are inevitably more subjective than those obtained by quantitative models based on numerical estimates. However, they represent a well-established and recognised framework to evaluate risk. In fact, the value of this method is precisely to allow a first and early identification of the relevant context-specific microbiological hazards and food products in the absence of food survey data but still based upon a systematic and logical reasoning of the evidence.

## 5. CONCLUSIONS

Good quality microbiological data on the presence of bacteria in food or epidemiological data on the frequency and the likely source of foodborne disease in the population is often lacking, particularly in LMICs. Even in the absence of such data, it is possible to systematically rank pathogens and food products according to the risk they pose, based on pathogen/products characteristics and context specific information including food regulations and risk profiles of food business operators. The approach applied to the dairy sector of Andhra Pradesh shown how the risk estimates that are generated, although of lower resolution if compared to data-driven risk assessments, can inform decision-making by identifying the pathogens and food products posing the higher risk for public health and the role of food business operators in shaping the risk.

## Data Availability

All data produced in the present work are contained in the manuscript

## AKNOWLEDGMENTS

This work was supported, by the Bill & Melinda Gates Foundation and the Foreign, Commonwealth and Development Office (FCDO) [Grant number: OPP1195676]. Under the grant conditions of the Foundation, a Creative Commons Attribution 4.0 Generic License has already been assigned to the Author Accepted Manuscript version that might arise from this submission. The research team would like to thank the individuals and organizations who generously shared their time, experience, and materials for the purposes of this project during the stakeholder meeting held in Andhra Pradesh. We are extremely greatefull to dr. Francesco Tranquillo for the thecnical support in the development of the Shiny app.

## Notes

### Competing Interest Statement

The authors have declared no competing interest.

### Funding Statement

This study was funded by the Bill & Melinda Gates Foundation

