## Supplementary Material #1 for "Microbiological risk ranking of foodborne pathogens and food products in scarce-data settings"

1. **Stakeholder workshop – milk supply chains in Andrah Pradesh**

In order to understand the dairy supply chain in AP and the quantity of milk flowing through different routes and actors along the value chain, a stakeholder workshop was held at NTR College of Veterinary Science, Gannavaram (AP), in February 2019. In addition to the researchers organizing and facilitating the workshop, the workshop was attended by thirteen participants, these included representatives of the Department of Animal Husbandry (N=2), Livestock Development Agency (N=2), Food Safety and Standards Authority of India (N=1), dairy cooperatives (N=3), private milk companies (N=2), dairy farmers (N=2) and a commercial raw milk retailer (N=1). A group exercise was conducted during which participants were separated into three groups with mixed composition to ensure representation of the different actors and asked to:

1. Map the routes and relative proportion of milk flowing through them from production to consumption.
2. List the various actors participating in each step of the chain.
3. List any government agencies, private companies or individuals influencing the behaviour of the actors at each step.

Information regarding typical and cultural consumption habits in relation to milk and dairy products and the current food safety regulatory framework were also collected. Following the group exercise, estimates of milk flows through the different routes were collectively discussed and a general flow chart of the milk flows from production to consumption was finalised with all participants until consensus was reached. A web-based application was developed to conveniently input and summarise the quantitative estimates collectively agreed from participants by means of a Sankey diagram. The app is freely available here: <https://mcrvc.shinyapps.io/riverflows/> and may be useful to those interested in representing, quantitatively, the complex flow of milk and dairy products through formal and informal channels that is common in low income settings.

While it is important to note that the intent of this exercise was to have a preliminary quantitative understanding of the amount of milk flowing through the different actors and not a precise estimate of the market shares represented by each channel, results of the mapping exercise were similar between the three groups working independently. This provides some reassurance of the accuracy of the chains being outlined and confidence that the quantitative estimates can be considered a realistic picture of the current milk supply chain in Andhra Pradesh.

**Stakeholder workshop – milk supply chains in Andrah Pradesh: Outputs**

**Structure of the milk supply chain in Andhra Pradesh**

It was estimated that around 25% of the total milk produced in Andhra Pradesh is consumed within the households or neighbourhood (ScN); this milk is mostly produced by household farms. Of the remaining 75% of milk produced that is marketed outside the local area, the largest shares of the transacted milk flow through the local collection centres (CC), followed by the traditional milk collectors (MC) and intermediary traders/private agents (PA). From the collection centre, the next destination is primarily the dairy cooperative plants for both chilling (COOPch) and production (COOPpr) of a variety of dairy products. From here, the product is sold to food business operators (FO) such as restaurants and hotels, other food industries (FI), or directly marketed to consumers through supermarkets (SK) and street vendors (SCS). A small amount of milk is sold to consumers directly at the collection centre (DS). The milk collected by milk collectors (MC) is dispatched to a variety of actors within the dairy industry, around half of it is typically sold directly to consumers (DS), the remaining is taken to the chilling centre, often owned by a private dairy company (PVch) or intermediary chilling centre (INTch). A small quantity of milk (not chilled) is directly sold to small scale manufacturers (SSMnch). Most of the milk flowing through the unregulated informal channel is supplied by households and small-scale farms through direct sales within the neighbourhood, village-based collection centres, or milk collectors. The exception is intermediary traders/private agents who mainly supply the private dairy industries with milk from large organised farms. Bulk milk coolers are not usually present at village level, therefore the cold chain is usually absent from milking upon delivery to the chilling centres from either the village-based collection centres or milk collectors. Intermediary traders/private agents mainly collect milk from organised farms (OF) and small-scale farms (SSF) and most of the milk is taken to the chilling centre of a private dairy industry (PVch). Only a small amount goes to small-scale manufacturers (SSMnch) and intermediary chilling centres (INTch). Small-scale manufacturers (SSMpr) mostly supply milk collectors (MC) or private companies (PVch) and, to a lesser extent, intermediaries (INTch). The main marketing channel for small-scale manufactures are street vendors (SCS) followed by supermarkets (SK) and direct sale to consumers (C). Private dairy industries are mainly supplied by their own chilling centre (PVch) storing the milk collected by intermediary traders/private agents (PA). Private industries may also process a small amount of milk from intermediary chilling centres (INTch). Market channels include other food industries (FI), food business operators (FO) or direct sale to street vendors (SCS) and supermarkets (SK).

**Regulatory framework**

Food safety and food regulation in India is overseen by the Food Safety and Standards Authority of India (FSSAI) a statutory body established under the preamble of the Food Safety Standards (FSS) Act, 2006 that also lays down the licencing and food safety requirements for food businesses (FSSAI, 2006). The type of registration/licence to be obtained by the FBOs is based upon the turnover, size and nature of the business. The Basic FSSAI Registration is issued by the State Government and is meant for FBOs such as processing units, milkman local shops and vendors of which production capacity does not exceed 100 L of milk/day or handling up to 500 L milk/day, these FBOs need to obtain registration with a self-attested declaration of adherence to hygiene and safety requirements. The State FSSAI License, issued by the State Government is meant for dairy processing units handling up to 50,000 L/day and operating in one state only while the Central FSSAI License is issued by the Central Government for processing units handling >50,000 L/day or with operations in more than one state. These FBOs are required to demonstrate compliance with Good Manufacturing Practices (GMP) and Cleaning-In-Place (CIP), regular microbiological testing “*as frequently as required on the basis of historical data and risk assessment*” and having in place recall plans (FSSAI, 2011). Despite having a legal framework in place, the effective enforcement of legislation in the current scenario is however challenging for a number of reasons ranging from FBOs (particularly small and medium business) awareness of FSSAI’s rules and regulations to difficulties in interpretation of the guidelines to compliance costs perceived as unsustainable and unjustified by many FBOs (Mehdi et al., 2019). Characterisation of the dairy system has identified different profiles of vendors (e.g. private dairy companies and cooperatives) with FBOs marketing milk and dairy products as industrially processed, packed and branded products being at the formal end of the formal-to-informal spectrum. These FBOs co-exist with FBOs selling dairy products without any license or registration (e.g. non-registered street vendors and mobile vendors) at the more informal end. Small-scale manufacturers also operate at different points of the spectrum. This category includes licensed and registered small to medium size FBOs that are formally operating within the regulated sector but in practice often do not meet the food safety standards and sanitary regulations in the processing and retail of dairy products.

**Context-specific information**

In terms of context-specific information, from discussion with stakeholders it appears that:

- Consumption of raw milk and raw milk products is negligible in AP.
- Consumers almost always boil milk prior to consumption; in some cases, even branded pasteurised or UHT milk.
- Milk flowing through the different supply chains might not always be stored at refrigeration temperature from the point of milking until delivery to the processing by the FBO, however, it is always pasteurised or sufficiently heat-treated prior to any manufacturing.
- FBOs in the dairy sector of AP are extremely diverse, what are normally defined as “formal” or “informal” often overlaps in a formal-to-informal spectrum of FBOs.
- Adherence to Good Manufacturing Practices (GMP), Cleaning-In-Place (CIP) and food safety Regulations in general is generally very good for FBOs in the formal end of the spectrum such as cooperatives and large private dairy companies. Moving towards the informal end of the spectrum, strict adherence to food safety regulations by FBOs progressively decreases until the very informal end represented by unregulated street vendors where basic food safety standards are largely disregarded.

2. **Description of the variables used in Multiple Correspondance Analysis**

*Initial Heat treatment; (IHT)****.*** For each product, IHT identifies whether a heat treatment deemed sufficient to kill the most heat-resistant vegetative pathogens (e.g. pasteurisation, sterilisation or equivalent) is performed at the beginning of the production process (yes/no). For some products (i.e. fermented products, see product-manufacturing table), this is the only heat treatment performed until packaging.

*Water activity; (a_w_).* Measured on a scale from 0 to 1, a_w_ is a measure of the water in a food matrix that is not bound to food components and available for microbial growth (Esener et al., 1981). In the absence of other inhibitory factors, the majority of microorganisms proliferate in foods with a_w_>0.95 while a_w_ of less than 0.86 is unfavourable for microbial growth of pathogenic bacteria (Demirci et al., 2020; Roos, 2003). Reducing a_w_ is an effective method of limiting the growth of pathogenic and spoilage organisms but should not be considered as a “kill” step (Beuchat et al., 2013). Products are categorised by a_w_ as: High (a_w_>0.95) assumed as the ideal for growth of all foodborne pathogens; Medium (0.86<a_w_<0.95), still favourable for most pathogens such as *Salmonella* spp. *E. coli* or *S. aureus* and Low (a_w_ <0.86), unfavourable for growth of all pathogenic bacteria (Majumdar et al., 2018; Roos, 2003).

*pH & Starter culture.* The pH is one of the key intrinsic factor capable of shaping the microbial metabolism in different ways, from the control of the bioavailability of nutrients to the activity of extracellular enzymes (Jin & Kirk, 2018). With a physiological pH value of about 6.8, milk is an ideal growth medium for most foodborne pathogens (Jay et al., 2005). The acidic pH of Indian dairy products mainly result from heat-acid coagulation as in the case of paneer or chhana, or bacterial fermentation for yoghurt or dahi (see product-manufacturing table). Although fermented products in general have an historical reputation of “safe” foods (Adams & Mitchell, 2002), fermented or acid-coagulated *dairy* products are considered as low-acid (i.e. pH above 4.6) foods. Hence, even though bacteria have narrow optimal pH ranges for growth as compared to yeasts and moulds (Demirci et al., 2020) the vast majority of foodborne pathogens are neutrophiles and growth is not inhibited at a pH value within up to two units from the neutral pH of 7. As such, the low acidic environment characterising some dairy products should not be considered as a factor preventing microbial growth.

Fermented dairy products on the other hand, are characterised by the addition of a starter culture, lactic acid bacteria (LABs) (Marth & Steele, 2001), after pasteurisation or equivalent heat treatment (see product manufacturing table) to ensure only LABs proliferate. LABs accelerate the production of lactic acid from the fermentation of sugars and secrete metabolites such as organic acids, hydrogen peroxide, and bacteriocins resulting in adverse effects on foodborne pathogens (Gao et al., 2019; Özogul & Hamed, 2018), also by means of suppression of pathogens' gene expression (Melo et al., 2016; Miranda et al., 2018). Furthermore, the presence of LABs in food represents a competition for available nutrients between foodborne pathogens. These factors combine to make the environment of fermented products generally unfavourable for growth of foodborne pathogens; for these reasons, dairy products were classified considering the addition of a starter culture (Starter=Yes/No) rather than merely the pH.

*Final Heat Treatment; (FHT).* For each product, FHT identifies whether a heat treatment that is deemed sufficient to kill the most heat-resistant vegetative pathogens (e.g. pasteurisation, sterilisation, prolonged cooking or equivalent) is performed as the final step of the production process (yes/no), a final heat treatment is considered effective in eliminating the bacteria that might have contaminated the product as a result of cross-contamination during processing.
