## Supplementary Material #2 for "Microbiological risk ranking of foodborne pathogens and food products in scarce-data settings"

RISK PROFILES

### *Aeromonas* spp.

| Shed in milk | No |
| --- | --- |
| Milk contaminant | Yes |
| Main source of indirect contamination | Water |
| Survive pasteurization | No |
| Severity of illness | Serious |
| T°C range (growth) | 4-45 C° |
| T°C optimum (growth) | 28-38 C° |
| pH range (growth) | 6-8.8 |
| pH Optimum (Range) | 6.5-7.5 |
| Aw | optimum: 0.991-0.986 + 1-2%NaCl |

*Aeromonas spp.* are ubiquitous and occur worldwide with water being considered as the main source of human infections (Daskalov, 2006; Verbyla & Rousselot, 2018). The predominant manifestation of *Aeromonas* infection in humans is diarrhoea, typically self-limiting. Bacteraemia and wound infection can happen in patients with underlying disease or healthy individuals following trauma and contact with contaminated water or soil (Figueras & Beaz-Hidalgo, 2015). Different epidemiological studies have associated *Aeromonas* infection with traveller’s diarrhoea in adults (Hanninen et al., 1995; Svenungsson et al., 2000; Vila et al., 2003; Yamada et al., 1997), however, human cases are typically sporadic and reporting is not mandatory; considering the self-limiting nature of the disease, it is reasonable to hypothesize that many cases go underreported. To our knowledge there has been only one large outbreak involving 2170 cases in which *Aeromonas* was identified as the etiologic agent (Hofer et al., 2006) but the source of infection was not identified. The microorganism has been isolated from a variety of food, from meat and edible organs of sheep and poultry to fish and seafood, raw milk, red meats and pork and beef. However, *Aeromonas* remains more frequently isolated from treated and untreated water and animals associated with water such as fish and shellfish (Igbinosa et al., 2012; Pessoa et al., 2019). Foods of animal origin, particularly fish and shellfish, may thus play a relevant role in the transmission of the *Aeromonas* spp. from food or animals to humans and faecal contamination/cross contamination with infected water in this case seems to be the major source of contamination (Ceylan et al., 2009; Verbyla & Rousselot, 2018). From the few studies investigating the occurrence of *Aeromonas spp.* in food samples in India, it seems that milk and dairy products are not to be considered as the food products posing the greater risk for the presence of the bacteria as compared to other foodstuff (Kumar et al., 2000; Manna et al., 2013). In general, from the few minor outbreaks for which an epidemiological link could be established, it seems that the food route has a marginal contribution to the global burden of disease if compared to exposure to contaminated water (Altwegg et al., 1991; Krovacek et al., 1995; Vally et al., 2004). It should also be noted that considering the clinical symptoms of *Aeromonas* spp. infection, the most severe forms (i.e. bacteraemia and wound) are unlikely to occur via ingestion of the bacteria for which the infectious dose seems to be high and >10^9^ CFU (Rusin et al., 1997). At present, there are not ISO standards for microbiological examination of foods for *Aeromonas spp.* for analysis of water and food samples it is necessary to refer to the culture media and protocols that can be found in literature ((Verbyla & Rousselot, 2018) and references therein).

### *Bacillus cereus*

| Shed in milk | No |
| --- | --- |
| Milk contaminant | Yes |
| Main source of indirect contamination | Soil, faeces, equipment, feed |
| Survive pasteurization | yes (fast-germinating spores) |
| Severity of illness | Moderate |
| T°C range (growth) | 4-55 C° |
| T°C optimum (growth) | 30-40 C° |
| T°C range (toxin production) | 12-37 C° Emetic toxins, 10-43 C° Diarrhoeal toxins |
| T°C optimum (toxin production) | 12-15 C° Emetic toxins, 32 C° Diarrhoeal toxins |
| pH range (growth) | 5-8.8 |
| pH Optimum (Range) | 6-7 |
| Aw | 0.93 |

Mainly two types of disease syndromes are caused by *B. cereus*. Emetic illness and Diarrhoeal illness. Emetic illness is mediated by highly stable toxins *pre-formed* in food withstanding passage through human stomach and causing vomiting after 1 to 6h from ingestion. In emetic type of illness, the dose is about 10^5^–10^8^ cells per gram of food in order to produce sufficient toxin (Gillespie & Hawkey, 2006).

Diarrhoeal illness is instead mediated by heat labile enterotoxins produced during growth of vegetative cells in the small intestine of the host and the infective dose is 10^4^–10^9^ cells per gram of food (Gillespie & Hawkey, 2006). These enterotoxins are normally destroyed by passage through stomach, hence, the risk is in this case posed by *B. cereus* itself colonising the intestine. *B. cereus* constitutes ~90% of the paddy soil bacteria and is considered a common contaminant of raw milk and dairy products (Tewari & Abdullah, 2015). Considering the infectious doses of the two syndromes, any food containing >10^3^ CFU/g should be considered as unsafe (Granum & Lund, 1997).

*B. cereus* is estimated to be one of the mayor foodborne pathogen worldwide (Scallan et al., 2011) but because of the typical short and mild course of disease, and the fact that symptoms are not easily distinguishable from intoxications caused by other foodborne pathogens (e.g. *S. aureus*), *B. cereus* infection are thought to be largely under-reported.

Contamination of pasteurized milk is typically attributed to the presence of pasteurisation-resistant spores in raw milk; the role of processing equipment as a reservoir for *B. cereus* milk recontamination is also well documented (Tewari & Abdullah, 2015). Several studies from different parts of India have identified the presence of *B. cereus* in various food and food products including milk (Chopra et al., 1980; Kumari & Kalimuddin, 2004; Kumari & Sarkar, 2014; Sharma et al., 2003; V. K. Singh et al., 2015). However, the vast majority of the *B. cereus* outbreaks were mostly associated with types of food different from milk and dairy products, in particular, products that are typically prepared in large amounts and stored inadequately (Bennett et al., 2013; Lentz et al., 2018; Stenfors Arnesen et al., 2008; Zhou et al., 2014).

Most *B. cereus* strains exhibit lipolytic and proteolytic activity resulting in product spoilage at temperatures greater than 7C° for long periods. In addition, the pathogen poorly competitive and is rapidly outgrown by gram-negative psychrotrophs at refrigeration temperatures. For this reason, milk and dairy products that are kept at refrigerated conditions normally spoil before *B. cereus* contamination is sufficient to cause illness. In fact, both syndromes described above are mainly consequence of the fact that *B. cereus* spores can survive heat treatments and under improper storage conditions, the spores germinate and the vegetative cells quickly multiply in food and/or produce high levels of toxin depending on the strain(s) being present (Wijnands, 2008).

When considering milk and dairy products, the following events should take place for the outbreak to occur: (i) the (unpasteurized) product contains B. cereus spores; (ii) the product is pasteurised (competitors removed) and re-contaminated before or during packaging (competitors not reintroduced) and (iii) the product is stored at temperatures allowing the spores to germinate. This is probably the reason why reported *B. cereus* outbreaks involving dairy products have been rare (Christiansson, 2011).

Methods to enumerate and detect *B. cereus* group are available (ISO 7932, ISO 21871), but it is important to be aware that they give no indication of the ability of the bacteria to produce toxins and are inadequate for the differentiation of *B. cereus sensu stricto* from closely related *B. cereus* group species that have not been associated with illness. Methods for detection of toxins and strain characterization for the production of toxins exist but are not standardised and not used routinely by industry.

### *Brucella* spp.

| Shed in milk | Yes |
| --- | --- |
| Milk contaminant | Yes |
| Main source of indirect contamination | Airborne, contact with infected animals. |
| Survive pasteurization | No |
| Severity of illness | Serious |
| facultative intracellular microorganism |  |

Genus Brucella includes: *B. abortus,* *B. melitensis,* *B. suis*, *B. ovis*, *B. neotomae* and *B. canis.* Brucellosis is repeatedly reported as one of the most important neglected zoonoses globally (WHO) including India ((Smits & Kadri, 2005) and references therein) with the most invasive species for human being, in descending order: *B. melitensis,* followed by *B. suis,* *B. abortus* and *B. canis (Corbel, 2006).*

The disease occurs worldwide, except in countries where bovine brucellosis (*B. abortus*) has been eradicated (eradication defined as absence of any reported cases for at least five years). The severity of the disease in humans is mainly determined by the Brucella specie to which one is exposed which in turn is significantly influenced by the animal host. In fact, while some cross-species infections occur, especially with *B. melitensis*, Brucellae are typically host-specific (Corbel, 2006). The main route of infection for ruminants is the oral mucosa and while *B. abortus* typically infects cattle *B. melitensis* and *B. ovis*, are the most common species infecting sheep and goats.

In cattle, the main symptom is infertility and abortion (premature or full term birth of dead or weak calves) usually taking place in the second half of gestation with retention of placenta and metritis (Poester et al., 2013). *B. melitensis* infection in sheep and goats affects the placenta and fetus with the main symptoms in pregnant goats being late gestation abortion (Poester et al., 2013). However, animals can develop self-limiting infections becoming asymptomatic latent carriers and potential shedders. In infected animals, the pathogen localizes in the supra-mammary lymph nodes and mammary glands, therefore, pathogen is secreted continuously or intermittently with milk.

Zoonotic transmission from infected animals to humans is either direct or indirect. Direct transmission is mediated by close contact with infected animals through respiratory, conjunctival and cutaneous routes or direct contact with the placenta, fetus and fetal fluids.

Brucellae aerosolize quickly, and through the respiratory route it is believed that a dose as little as 10 CFU if enough for infection (*B. melitensis*, *B. suis,* *B. abortus* being even listed as potential bio-weapons by the Centres for Disease Control and Prevention in the USA) (Seleem et al., 2010). The infection dose from oral (ingestion) route is however unknown.

Indirect transmission to humans is food-borne and mainly associated with consumption of raw milk and dairy products made from unpasteurized milk of sheep and goats (Dadar et al., 2019). Symptoms in humans are non-specific and can be confused with those of influenza, the WHO recommend the following definition for clinical human cases: *“An illness characterized by acute or insidious onset, continued, intermittent or irregular fever of variable duration, profuse sweating, particularly at night, fatigue, anorexia, weight loss, headache, arthralgia, and generalized aching. Local infection of organs may occur.”*

*B. abortus* is the most widespread cause of infection, often sub-clinical in humans, is usually less severe than that caused by *B. melitensis* or *B. suis (Acha & Szyfres, 2003)*. *B. ovis* is of little significance in relation to human disease. Brucella species are facultative intracellular pathogens that can survive and multiply within phagocytic cells of the host. Survival in milk and milk products has been observed to decline as the storage temperature increases or the pH decrease. Conversely, high fat content of products might have protective effects (Salem, 1977).

Bacterial isolation is considered as the “gold standard” for specific diagnosis, however, *Brucella spp.* requires the use of biological safety level BSL-3 or BSL-2 with BSL-3 precautions (Chosewood & Wilson, 2009) for all manipulations of specimens and cultures*.* The conventional indirect methods to evaluate presence of Brucella contamination in serum or milk are Enzyme-Linked Immunosorbent Assay (ELISA), Milk Ring Test (MRT) and the Rose Bengal Test (RBT but it should be stressed that while useful for control and surveillance, these techniques do not measure the actual presence of the microorganism.

### *Campylobacter* spp.

| Shed in milk | No |
| --- | --- |
| Milk contaminant | Yes |
| Main source of indirect contamination | Faeces |
| Survive pasteurization | No |
| Severity of illness | Moderate |
| T°C range (growth) | 30-45°C (Microaerophilic conditions) |
| T°C optimum (growth) | 42-45 |
| T°C range (toxin production) | n.a. |
| T°C optimum (toxin production) | n.a. |
| pH range (growth) | 4.9-9.5 |
| pH Optimum (Range) | >4.6-? |
| Minimum a_w_ (for growth) | 0.99 |

With almost 1 in 10 people falling ill and 33 million of healthy life years lost every year due to campylobacteriosis, *Campylobacter* is a zoonotic pathogen of global relevance (Havelaar et al., 2015; WHO, 2018), the main route of transmission is believed to be foodborne, particularly via undercooked meat products or contaminated raw milk.

The two main pathogenic strains are *C. jejuni* and *C. coli* causing fever and enteritis in human. *Campylobacter* enteritis is acute inflammatory diarrhoea with clinical signs similar to other enterobacteriaceae such as salmonellosis or shigellosis. Although infection in human is not severe with complete recovery without treatment in few days, campylobacters cause the highest number of human gastroenteritis cases in developed economies and accounts for about half of the food-borne illnesses caused by bacteria worldwide.

The main environmental niche for this pathogen is the intestinal tract of all avian species, but Campylobacters are also part of the normal intestinal flora a variety of farm and companion animals amongst which cattle, sheep, goats, pigs cats and dogs (WHO, 2018). While the source of the majority of the *Campylobacter* spp. outbreaks have remained unknown, poultry and dairy products (raw milk) are the food categories most commonly identified as the causal source (CDC, 2019; EFSA, 2018).

The pathogen does not survive pasteurisation and require microaerophilic conditions for growth. Campylobacters do not multiply at temperatures below 30°C, meaning that the microbial load in food will not increase at typical room temperature of 20-25°C. Although unable to grow below 30°C, *Campylobacter* remains metabolically active and is motile at temperatures as low as 4°C. The pathogen is sensitive to heat treatments and destroyed by pasteurisation; cooking at 55-60°C for several minutes is also deemed sufficient for inactivation.

Mastitis caused by *Campylobacter* are rare (Skirrow, 1991) and infection from consumption of milk mainly results from faecal contamination. Amongst dairy products, raw milk or products made with unpasteurised or improperly pasteurised milk have been identified as the source of infection in the vast majority of the dairy-related outbreaks. Dose-response models for *Campylobacter jejuni* have estimated doses required for 50% illness (IllD50) as little as 1.7x10^2^ CFU (median value) from human challenge data but with significant differences by strains (Teunis et al., 2018).

### *Clostridium botulinum*

| Shed in milk | No |
| --- | --- |
| Milk contaminant | Yes |
| Main source of indirect contamination | Faeces, silage of poor quality |
| Survive pasteurization | yes (spores)  no (neurotoxin) |
| Severity of illness | Severe |
| T°C range (growth) | 10-48 C° (minimum 3.3 C° non-proteolytic strains) |
| T°C optimum (growth) | 35 |
| T°C range (toxin production) | // |
| T°C optimum (toxin production) | 35 |
| pH range (growth) | 4.6-8.9 |
| pH Optimum (Range) | >4.6-? |
| Minimum aw (for growth) | 0.95 |

Foodborne botulism is a rare but serious and potentially fatal disease. As the bacterium is strictly anaerobic, intoxication occurs when *C. botulinum* produces toxins in food prior to consumption. In fact, consuming low levels of botulinum spores per se rarely poses a health issue to individuals older than one year old, unless they are devoid of intestinal microflora; this is because natural defences in intestines that develop over time prevent germination and growth of the bacterium. Children and adults consume botulinum spores without adverse effect when eating foods such as honey, vegetables that grown in or near the ground (such as carrots and potatoes), or smoked fish. Similarly, a low numbers of clostridia spores in dried milk products or other dried foods do not present a health risk (Doyle & Glass, 2013). A problem could occur if the product is rehydrated and kept anaerobically at environmental temperature, this would allow the spores to germinate, grow, and produce neurotoxins. While spores of *C. botulinum* are heat-resistant, the toxins produced by bacteria are not thermostable (Weingart et al., 2010). Therefore, prerequisites for the intoxication form foodborne botulism are: (1) Milk contains *C. botulinum* spores or vegetative cells, (2) *C. botulinum* spores or vegetative cells survive food processing, (3) Storage conditions are such to enable *C. botulinum* spore germination, growth, and toxin production, and (4) neurotoxin-containing food is ingested without reheating.

Human cases are caused mostly by toxin types A, B, E, and F; exact lethal dose are uncertain but for type A, primate studies indicate a dose of 0.001 μg/kg of body weight as lethal from oral (ingestion) route (Arnon et al., 2001).

In contrast to intoxication, events leading to intestinal botulism include (1) ingestion of *C. botulinum* spores, and (2) altered or abnormal bacterial population in the gastro intestinal tract due to young age, heavy antibiotic treatment, or intestinal lesions, allowing spore germination, growth, and toxin synthesis.

*C. botulinum* is widely ubiquitously distributed in soils and aquatic sediments, and has been detected in almost all type of food of plant and animal origin. However, even though food may contain viable *C. botulinum* and satisfy the nutritional requirements for growth, if anaerobic conditions are not met, growth and toxin production will not take place. In addition, although a low pH does not degrade any pre-formed toxin, the pathogen does not grow in acidic conditions (pH < 4.6), and therefore, the toxin will not be formed in acidic foods. For these reasons, *C. botulinum* is a major concern when heat treatments are used to extend the shelf life of products that are then canned or packed under modified atmosphere. In this case, if spores are present, they can survive the heat treatment prior to packaging and the anaerobic conditions can promote growth and toxin production, especially if the food is subjected to temperature abuse. Not surprisingly, the source of foodborne botulism is often acidic, home-canned foods such as fruits, vegetables and fish. Multiplication of *C. botulinum* in silage and in the gastrointestinal tract of cattle with botulism has been reported, therefore, contamination of the farm environment and raw milk, and further transmission through the dairy chain, is possible (Bohnel & Gessler, 2013; Frank, 2013).

Although rare, large botulism outbreaks due to both commercial and home-prepared dairy products have been reported. Factors explaining these outbreaks include most importantly temperature abuse, but also unsafe formulation, inadequate fermentation, insufficient thermal processing, post-process contamination, and lack of adequate quality control for adjunct ingredients. The small number of outbreaks is probably explained by a low incidence of spores in milk due to infected cows, the presence of competitive bacteria, and growth-inhibitory combinations of intrinsic and extrinsic factors in cultured and processed dairy products.

There are not standardised methods for enumeration of *C. botulinum*. Detection of *C. botulinum* is based on screening for the toxin (in vivo test, ELISA, testing for enzyme activity) and detection of the bacteria by enrichment broth culture followed by detection of toxin and/or genes encoding neurotoxins by PCR.

### *Corynebacterium* spp.

| Shed in milk | Yes |
| --- | --- |
| Milk contaminant | Yes |
| Main source of indirect contamination | Human droplets |
| Survive pasteurization | No |
| Severity of illness | Serious |
| T°C range (growth) | 15-40 C° |
| T°C optimum (growth) | 37 C° |
| T°C range (toxin production) | // |
| T°C optimum (toxin production) | // |
| pH range (growth) |  |
| pH Optimum (Range) | 7.2-7.6 |
| Minimum a_w_ (for growth) |  |

Genus Corynebacterium comprises a diverse group of bacteria, including both animal and plant pathogens. *C. bovis* is one of the most important mastitis-causing microorganisms together with *S. aureus* (Underwood et al., 2015). Bacteria of this genus include also *C. ulcerans,* *C. pseudotuberculosis* and *C. diphtehriae*.

Reservoir of *C. diphtheriae* are humans and transmission is from person to person by intimate respiratory and direct contact (Hadfield et al., 2000). On the other hand, *C. ulcerans* and *C. pseudotuberculosis* are zoonotic infections, mainly associated with handling of infected dairy/companion animals or consumption of contaminated milk (Tiwari et al., 2008; Wagner et al., 2010); person-to-person transmission for these species has been hypothesized but not clearly documented (Konrad et al., 2015). *C. ulcerans* and *C. pseudotuberculosis* can harbour diphtheria *tox* genes and express diphtheria toxin for which, a lethal dose of < 0.1 µg/kg of body weight in humans has been estimated (Zakikhany & Efstratiou, 2012).

Very few studies have been conducted for the presence of *Corynebacterium spp.* in food.

### *Coxiella burnetii*

| Shed in milk | Yes |
| --- | --- |
| Milk contaminant | Yes |
| Main source of indirect contamination | Urine, faeces, vaginal mucus. |
| Survive pasteurization | No |
| Severity of illness | Serious |
| Obligate intracellular microorganism |  |

*Coxiella burnetii* is the foodborne pathogen responsible for Q fever in humans. Q fever in humans is primarily transmitted through inhalation of dust contaminated with infected animal birth products or secretions, such as the placenta (McDaniel et al., 2014). While there is evidence viable *C. burnetii* in milk (Loftis et al., 2010) and consumption of raw milk is considered as a suspected mode of transmission, (particularly from milk of animals with history of reproductive disorder). The role of milk as actual vehicle of transmission and source of infection in humans remains controversial (McDaniel et al., 2014).

While highly infectious through inhalation, experiments in which contaminated milk was fed to volunteers or prisoners were contradictory (Angelakis & Raoult, 2010) and the supporting evidence of *C. burnetii* being a significant pathogen causing clinical disease through ingestion of milk is weak to the point that some authors even challenged the role of *C. burnetii* as food-borne pathogen (Cerf & Condron, 2006).

*C. brunettii* outbreaks associated with windborne transmission or contact with animals are well documented (Brouqui et al., 2004; Hogerwerf et al., 2012; Tissot-Dupont et al., 2004; Wilson et al., 2010) and there seems to be little doubts that these are the main route of infection (Gale et al., 2015). As precautionary principle, it can be concluded that *C. burnetii* might pose a risk for consumers if present at high levels in milk and the milk is consumed as raw.

### *Cryptosporidium* spp.

| Shed in milk | No |
| --- | --- |
| Milk contaminant | Yes |
| Main source of indirect contamination | Water, faeces. |
| Survive pasteurization | No |
| Severity of illness | Severe |
| Obligate intracellular protozoan |  |

Cryptosporidium is an intestinal protozoan considered to be one of the most important food borne parasites globally (Bamaiyi & Redhuan, 2017). The most common symptoms of cryptosporidiosis are watery diarrhoea and stomach cramps, vomiting, nausea and fever. Infection is typically self-limiting, from few days to weeks unless the immune system is already compromised in which case symptoms can last for months. Indeed, cryptosporidiosis is the second biggest cause of infant diarrhoea and death in Africa and Asia (Striepen, 2013).

As a parasite excreted with faeces, transmission to humans occur through accidental ingestion of *Cryptosporidium* oocytes excreted from feces of animals or humans. Direct contact with cattle or consumption of food or water contaminated with cattle manure has been identified as the cause of many cryptosporidiosis outbreaks ((Xiao & Feng, 2008) and references therein). *Cryptosporidium* can survive the wastewater treatments and is not inactivated by the commonly used drinking water disinfectants (Zahedi et al., 2016); as the main causative agents of the majority (63%) of the waterborne protozoan parasitic outbreaks worldwide since 2004 (Baldursson & Karanis, 2011; Efstratiou et al., 2017), *Cryptosporidium* is undoubtedly a major public health concern for the global water industry. The median infective dose calculated by means of linear regression using human challenge data on healthy volunteers was 132 oocyst (DuPont et al., 1995) infections may occur with ingestion of as few as 30 or even 1 oocysts (Guerrant, 1997).

Infection in milking animals leads to diarrhoea and shedding in the faeces. Hence, milk might be contaminated via direct contact with faecal material during milking, contaminated water or infected workers. Oocysts can survive in wet/moist foods, and remain infective in most conditions for long periods at refrigeration temperature, even at pH ~4.0 (Dawson et al., 2004). However, the oocysts appear to be sensitive to heat, loosing infectivity rapidly after 1 min at >60°C (Rose, 1997), in fact, cryptosporidiosis outbreaks involving dairy products are mainly associated to consumption of unpasteurised milk.

While several studies conducted in different part of the world (including India with prevalence estimates ranging from 2.8% to 26% (Joute et al., 2016; Kashyap et al., 2019; Singh et al., 2018)) have investigated the prevalence of cryptosporidiosis in cattle (Ayele et al., 2018; Lefay et al., 2000; Nguyen et al., 2007; Santın et al., 2004), very few studies have been undertaken to determine the prevalence of *Cryptosporidium* oocysts in food.

Of those that are available, methodologies to isolate oocysts from samples are not consistent, together with methods of detection and viability assays. Methods for detection and enumeration of the existing numbers of oocysts that are likely to be present in the food material are complex and many steps are required (elution, concentration, purification and isolation) just to prepare the food sample for the detection step (Ahmed & Karanis, 2018).

### *Cronobacter (Enterobacter) sakazakii*

| Shed in milk | No |
| --- | --- |
| Milk contaminant | Yes |
| Main source of indirect contamination | Unknown |
| Survive pasteurization | No |
| Severity of illness | severe (infants) |
| T°C range (growth) | 6-45 C° |
| T°C optimum (growth) | 37-43 C° |
| T°C range (toxin production) | // |
| T°C optimum (toxin production) | // |
| pH range (growth) | 3.9-? |
| pH Optimum (Range) | 5-9 |
| Minimum a_w_ (for growth) | Survives in PIF (aw = 0.2) |

Enterobacter sakazakii (reclassified as *Cronobacter* spp. in 2007) is a gram-negative, non-spore-forming bacterium belonging to the Enterobacteriaceae family. The pathogen is considered to be ubiquitous and has been detected from a variety of food and environments (Baumgartner et al., 2009; Jaradat et al., 2009; Putthana et al., 2012; N. Singh et al., 2015) including milk powder, powdered infant formula (PIF) and milk protein-producing facilities where the presence of *C. sakazakii* is common and well documented ((Jacobs et al., 2011) and references therein). *C. sakazakii* can form biofilms promoting adherence and environmental stress resistance (Grimm et al., 2008; Lehner et al., 2005), this is probably the main reason why the bacteria is commonly detected on food contact surfaces and processing environments. While the main reservoir for *Cronobacter spp.* remains unknown, PIFs are repeatedly identified as the main vehicle of infection with a lethal dose tentatively estimated as ranging between 10^3^ and 10^8^ (WHO/FAO, 2004). Considering the high (from 40% to 80%) reported mortality rate and severe sequelae (i.e. necrotizing enterocolitis, bacteraemia, and meningitis) associated with the illness (Nazarowec-White & Farber, 1997), significant efforts have been devoted to assess the food safety risk of *C. sakazakii* in PIFs (WHO/FAO, 2004).

*C. sakazakii* is a thermotolerant organism but not survive pasteurisation; although PIFs are pasteurised and *C. sakazakii* does not grow on dry substrates, the microorganism can survive for a long time on dry environments as compared to other enterobacteriaceae (Breeuwer et al., 2003). This provide competitive advantages against other bacteria and increases the risk of post-pasteurization contamination of the finished product. Similarly to *B. cereus*, *C. sakazakii* can grow in reconstituted milk powders and PIFs if stored above 5°C for a sufficient time and multiply very rapidly at room temperatures. Contamination and subsequent growth may occur during reconstitution, preparation, handling and filling, via raw materials and in particular heat sensitive nutrients, (e.g. vitamins, minerals etc.) added after pasteurisation and/or cross-contamination from the processing environment.

### *Escherichia coli* O157:H7

| Shed in milk | No |
| --- | --- |
| Milk contaminant | Yes |
| Main source of indirect contamination | Faeces |
| Survive pasteurization | No |
| Severity of illness | Severe |
| T°C range (growth) | 6-45 C° |
| T°C optimum (growth) | 37 C° |
| T°C range (toxin production) | ? |
| T°C optimum (toxin production) | ? |
| pH range (growth) | 3.9-? |
| pH Optimum (Range) | 4.4-8.5 |
| Minimum a_w_ (for growth) | 0.95 |

Part of enterobacteriaceae *Escherichia coli* is normally present in the intestinal microflora of humans and other animals (Gyles, 2007; La Ragione et al., 2009). While most strains are harmless, some are pathogenic to humans and can cause severe life threating infections. The Shiga toxins-producing EHEC serotype *E.coli 0157:H7* (STEC) in particular, is especially virulent, zoonotic and responsible for the majority of the pathogenic *E.coli* foodborne cases worldwide. The pathogen is naturally present is the intestines of cattle (Lim et al., 2010) and is shed continuously or intermittently with faeces at heterogeneous doses (Lim et al., 2010; Robinson et al., 2004) creating the potential for cross-contamination of beef, milk and dairy products and vegetables through cattle manure (Mora et al., 2007).

While cattle are asymptomatic, infection in humans is typically characterised by diarrhoea like symptoms. Haemorrhagic colitis, an acute illness is characterised by severe abdominal pain and bloody diarrhoea. About 5% of the haemorrhagic colitis victims may develop haemolytic uremic syndrome (HUS) as a results from Shiga toxins (Stxs) produced by the bacteria in the intestine (Gyles, 2007) with a fatality rate of around 5% (Ameer et al., 2021).

Infection with pathogenic *E. coli* is a cause of significant morbidity and mortality worldwide with beef and dairy ranking in first and third position respectively in terms of source of infection from outbreak surveillance data (FAO/WHO, 2019). As natural part of the intestinal microflora of ruminants, foods derived from these animals might become contaminated as consequence of faecal contamination during processing.

Prevalence of *E. coli O157:H7* in dairy products has been determined in a limited number of studies in different areas of the world with the vast majority involving raw milk or cheese produced with raw milk. These include India (Neher et al., 2015; Sethulekshmi & Latha, 2016; Vanitha et al., 2018) where it was also demonstrated how *E. coli O157:H7* could grow and survive during manufacturing of paneer if made with contaminated milk (Wahi et al., 2006). *E. coli* O157:H7 is extremely acid resistant, which contributes to the low infectious dose for humans estimated to be fewer than 100 CFU and possibly even as low as 10 (Constable et al., 2017).

### *Leptospira*

| Shed in milk | No |
| --- | --- |
| Milk contaminant | Yes |
| Main source of indirect contamination | Urine, soil, water |
| Survive pasteurization | No |
| Severity of illness | Severe |
| T°C range (growth) | 10-42 °C |
| T°C optimum (growth) | 28-30 °C |
| pH range (growth) | 6.9–7.4 |
| pH Optimum (Range) | 7.2-7.6 |
| Minimum a_w_ (for growth) | ? |

Leptospirosis is a contagious disease which infects both animals and humans, the bacteria multiply in the kidneys of animals and are shed with urine. Considered as an occupational hazard for farmers, veterinarians or abattoir workers, human infection results from exposure to infected urine of carrier mammals, either directly or via contamination of soil or water (Bharti et al., 2003).

The disease is often difficult to distinguish from other causes of febrile illness, severe forms of Leptospirosis occur in 5-15% of the clinical cases and of these the fatality rate is estimated to be around 50% (Torgerson et al., 2015).

While infection via oral route cannot be excluded, Leptospirosis caused by drinking water has not been as frequently reported as percutaneous infection. Hence, the main route of transmission remains the contact of the damaged skin and mucous membranes with water, damp soil or vegetation contaminated, in particular, with rodent urine. In fact, most of the reported *Leptospira* outbreaks occurred during flooding after heavy rainfall when the spread of the organism was facilitated as a consequence of the proliferation of rodents shedding large amounts of microorganisms in their urine. Reasons for the minor role of the oral route in the burden of Leptospirosis seems to be the efficiency of the natural defences (i.e. human saliva and gastric acid) against oral infection (Asoh et al., 2014).

In general, pathogenic leptospira survives in many environments for a long period of time, lasting months or years, allowing reinfection of animal hosts (Andre-Fontaine et al., 2015; Picardeau, 2017). The environmental conditions for survival are, however, narrow and the bacteria tend not to be a significant problem ex-vivo as opposite to other bacteria or viruses. In raw milk artificially contaminated with *Leptospira*, the pathogen was not detected by microscopic observations carried out weekly during the trial period. Milk seems therefore not to be a suitable media for the pathogen (Fratini et al., 2016).

### *Listeria monocytogenes*

| Shed in milk | Yes (rare) |
| --- | --- |
| Milk contaminant | Yes |
| Main source of indirect contamination | Environment |
| Survive pasteurization | No |
| Severity of illness | Severe |
| T°C range (growth) | -1-45 °C |
| T°C optimum (growth) | 37 °C |
| pH range (growth) | 4.4–9.4 |
| pH Optimum (Range) | 7 |
| Minimum a_w_ (for growth) | 0.92 |

Food-borne exposure is the primary route of transmission for listeriosis, a relatively rare disease (0.1 to 10 cases per 1 million people/year depending on the country) but characterised by a case-fatality rate of about 20% (CDC, 2021a) with nearly one-quarter of pregnancy-associated cases result in fetal loss or death of the new-born. For these reasons, listeriosis is considered a primary public health concern. The FAO estimated the infectious dose of listeriosis as between 10^7^–10^9^ CFU and 10^5^–10^7^ for the normal and high risk population respectively (FAO/WHO, 2004).

*L. monocytogenes* is ubiquitous in nature and milk and other dairy products can become contaminated at many stages along the food chain, from the faeces of milking animals, from the food manufacturing environment (Tabit, 2018), or, in rare cases from udder infection (Hunt et al., 2012; Winter et al., 2004).

The pathogen can grow in a broad range of temperature and pH (George et al., 1988) and food fermentations involving gradual lowering of pH could lead to acid adaptation of *L. monocytogenes (Saklani-Jusforgues et al., 2000)*. In addition, when compared to other food-borne pathogen, *L. monocytogenes* has the ability to multiply at aw values as low as 0.90. *L. monocytogenes* is also tolerant to salt (Liu et al., 2005) and can grow in NaCl concentrations up to 10% (McClure et al., 1989). These characteristics make virtually any dairy product as the ideal environment for *L. monocytogenes* growth and survival.

In fact, outbreaks of invasive listeriosis (the most severe form of listeria illness) have been linked to soft cheeses; soft, semisoft and mould-ripened cheeses; pasteurised chocolate flavoured milk; pasteurised and unpasteurised milk; butter; unpasteurised ice cream; ricotta cheese; goat, sheep and feta (CDC, 2021b; Lundén et al., 2004). Few studies have reported the occurrence of *L. monocytogenes* in dairy products in India (Aurora et al., 2008; Dhanashree et al., 2003; Kalorey et al., 2008; Soni et al., 2013).

### *Mycobacterium bovis*

| Shed in milk | Yes |
| --- | --- |
| Milk contaminant | Yes |
| Main source of indirect contamination | Faeces |
| Survive pasteurization | No |
| Severity of illness | *Severe* |
| Obligate intracellular organism |  |

*Mycobacterium bovis* is a related member of the *Mycobacterium tuberculosis* Complex, the causative agent of human tuberculosis (TB) a major cause of illness and mortality worldwide. Clinical symptoms of gastrointestinal infection include: fever, weight loss, abdominal pain, diarrhoea or constipation. Symptoms may last for years and death may result, particularly in case of compromised immune system or reactivation of latent infection occurring in old age, when resistance to mycobacterial infection is lowered.

The disease is primarily caused by *M. tuberculosis* which is usually transmitted through the respiratory route by close contact and inhalation of infected aerosols. Zoonotic tuberculosis due to *M. bovis* is mainly indirect through consumption of raw milk, dairy products made with raw milk or meat from infected cows (WHO/FAO/OIE, 2017). Pasteurisation of milk is deemed sufficient to eliminate the pathogen and in industrialized countries, animal TB control and elimination programs, together with milk pasteurization, have drastically reduced the incidence of disease caused by *M. bovis* in both cattle and humans (Kleeberg, 1984).

Infected livestock herds are now uncommon in Europe, Canada, the United States, New Zealand and some other locations but bovine tuberculosis is still common or relatively common in cattle in parts of Africa, Asia, the Middle East and Latin America (CFSPH, 2019).

Airborne and direct contact infections continues to occur among meat industry and slaughterhouse workers in regions where the infection is still prevalent in cattle and in middle- and low-income settings where pasteurization is less widely implemented. It seems that infection (animal-experimental conditions) via the oral route requires thousands or millions of organisms as compared to less than 10 through inhalation (de la Rua-Domenech, 2006; Thoen et al., 2014). *M. bovis* is an intracellular pathogens, capable of growing and multiply only inside macrophage cells; the slow growth combined with the short shelf life of the dairy products makes *M. bovis* unlikely to grow to any significant extent in food during production, processing, distribution and storage. The main risk of infection within the dairy supply chain seems thus to be confined to consumption of highly contaminated raw milk.

As for the *Mycobacterium avium* subsp. *paratuberculosis*, also *M. bovis* is slow growing (duplication time ~20h) and as an obligate aerobes microorganism, difficult to culture as it does not grow on ordinary microbiological media (FDA, 2012).

### *Salmonella* spp.

| Shed in milk | Yes (rare) |
| --- | --- |
| Milk contaminant | Yes |
| Main source of indirect contamination | Faeces |
| Survive pasteurization | No |
| Severity of illness | Severe |
| T°C range (growth) | 5.2-46°C |
| T°C optimum (growth) | 35-43 °C |
| pH range (growth) | 3.8–9.5 |
| pH Optimum (Range) | 7-7.5 |
| Minimum a_w_ (for growth) | 0.94 |

Salmonella is a Gram-negative genus belonging to the Enterobacteriaceae family. Within the two species: *Salmonella bongori* and *Samonella enterica,* over 2500 serovars have been identified so far. Few serovars are host specific with *Salmonella enterica* serotype Dublin being specific for cattle and *Salmonella enterica* serotype Choleraesuis for pigs, however, most serotypes reported as causative agent of human infections are present in a wide range of hosts. Salmonellosis is one of the most commonly reported enteric illnesses worldwide (WHO, 2015), of particular public health relevance are *S. enterica* serovars Typhimurium and Enteritidis, the two most common serotypes transmitted from animals to humans in most parts of the world.

The severity of *Salmonella* infections varies depending on the serovars and the underlying health status and age of the human host (Eng et al., 2015). Infection through the oral route is usually self-limiting and can range from having no effect, to colonisation of the gastrointestinal tract without symptoms, or colonisation with the typical symptoms of acute gastroenteritis. The infectious dose is estimated to be about 10^5^ CFU (Kenneth J. R et al., 2020), severe cases of diarrhoea might lead to significant dehydration in which case medical intervention is needed with intravenous fluid replacement (Baron, 1996). In a small number of cases, particularly in presence of serious underlying disease, infection can be life-treating (Kennedy et al., 2004).

Salmonellae are transmitted by the faecal-oral route, known routes of transmission include person-to-person (particularly for *S. Typhi* and *S. Paratyphi* for which humans are the only known reservoirs (Connor & Schwartz, 2005)), food, untreated drinking water and direct contact with infected animals. Primary reservoir of *Salmonella* is the intestinal tract of animals, infected animals can shed *Salmonella* with faeces and the occurrence of positive animals at the time of slaughter might lead to cross contamination with faeces during processing. Raw meat products are in fact amongst those more frequently associated with the presence of the pathogen, together with eggs (EFSA, 2019).

### *Shigella* spp.

| Shed in milk | No |
| --- | --- |
| Milk contaminant | Yes |
| Main source of indirect contamination | Water, faeces |
| Survive pasteurization | No |
| Severity of illness | Serious |
| T°C range (growth) | 6-48 °C |
| T°C optimum (growth) | 20 °C |
| pH range (growth) | 4.5-9.3 |
| pH Optimum (Range) | 6-8 |
| Minimum a_w_ (for growth) | 0.96 |

*Shigella* is a genus of the Enterobacteriaceae family including four species: *S. dysenteriae, S. flexneri, S. boydii,* and *S. sonnei* with several serotypes being described for the first three species.

Although shigellosis can be asymptomatic, common clinical signs of human shigellosis include diarrhoea (sometimes bloody), fever, stomach pain and the feeling of the need to pass stool even when the bowels are empty. Symptoms usually last 5 to 7 days, but some people may experience symptoms anywhere from a few days to 4 or more weeks (CDC, 2017).

The only known natural reservoirs of *Shigella* are humans and large primates (Dekker & Frank, 2015). As for the other intestinal pathogens, transmission of *Shigella* is via the faecal-oral route including person-to-person or consumption of water or food contaminated with feces from infected individuals. As few as 10 organisms can cause infection (DuPont et al., 1989), this makes shigellosis as very easily transmitted.

Not surprisingly, shigellosis outbreaks have occurred more commonly under conditions that facilitated the spread of bacteria through the faecal-oral route. Inadequate treatment of water used for drinking or food preparation, inadequately disinfected swimming pool or recreational water contaminated with human faeces have been identified as the main potential sources of transmission of shigellae.

Contamination of food is generally the result of poor personal hygiene of food handlers or in case of vegetables, use of human fertilisers (Lightfoot, 2003; WHO, 2005).

### *Staphylococcus aureus*

| Shed in milk | Yes |
| --- | --- |
| Milk contaminant | Yes |
| Main source of indirect contamination | Environmental, humans |
| Survive pasteurization | No (Bacteria), Yes (Toxins) |
| Severity of illness | Moderate |
| T°C range (growth) | 7-48 °C |
| T°C optimum (growth) | 37 °C |
| T°C range (toxin production) | 10-48°C |
| T°C optimum (toxin production) | 40-45°C |
| pH range (growth) | 4-10 |
| pH Optimum (Range) | 4.5-9.6 for toxin production |
| Minimum a_w_ (for growth) | 0.86 (for toxin production) |

*S. aureus* is among the leading causes of foodborne bacterial intoxications worldwide (Fetsch & Johler, 2018), it is an ubiquitous microorganism that can be detected on mucous membranes and skin of most animals, including all dairy animals and humans (Wertheim et al., 2005). *S. aureus* is also one of the most important cause of mastitis in cows and can be shed in milk in high amounts.

Staphylococcal food-borne illnesses are caused by the ingestion of food that contains preformed toxins produced by coagulase positive *S. aureus* strains. So far, more than 20 staphylococcal enterotoxins have been identified (Pinchuk et al., 2010). The production of enterotoxins is dependent on de novo synthesis within the cell and some of them seems to be produced only as a function of a “quorum-sensing” mechanism (Schelin et al., 2011) requiring the pathogen to be present above a certain amount for the enterotoxin to be produced.

While the pathogen is readily killed at cooking and pasteurisation temperatures, enterotoxins are extremely heat-resistant and their potency can only be gradually decreased by prolonged boiling or autoclaving (Ortega et al., 2010). For these reasons, control of growth of this organism prior to heat treatment is essential.

Symptoms generally appear around 3 hours after ingestion and include nausea, vomiting, abdominal cramps and diarrhoea. Symptoms are self-limiting with full recovery between 1-3 days requiring no medical treatment (Le Loir et al., 2003). Despite being one of the most common type of food-borne diseases worldwide, staphylococcal food poisoning goes often under-reported.

Many different foods are a good medium for *S. aureus* survival and growth, indeed, a variety of food have been implicated in *S. aureus* outbreaks, including dairy products such as milk and cream, butter and cheese or cream-filled pastries, ham, sausages, canned meat, salads, or ready-to eat sandwiches. The main sources of post-heat treatment contamination are humans as a consequence of handling or coughing and sneezing, however, for food such as raw meat, sausages, raw milk, and raw milk cheese, contaminations from animal origins are more frequent and due to animal carriage or contaminated milk (Le Loir et al., 2003).

### *Streptococcus* spp.

| Shed in milk | Yes |
| --- | --- |
| Milk contaminant | Yes |
| Main source of indirect contamination |  |
| Survive pasteurization | No |
| Severity of illness | serious |
| T°C range (growth) | 10-45 °C |
| T°C optimum (growth) | 37 °C |
| T°C range (toxin production) | ? |
| T°C optimum (toxin production) | ? |
| pH range (growth) | 4.8-9.3 |
| pH Optimum (Range) | 7 |
| a_w_ | ? |

Streptococci are Gram-positive cocci in the family Streptococcaceae, some species of *Streptococcus* are part of the normal flora in both humans and animals but few species with reservoirs in animals have been proven to be zoonotic. In particular, *S. equi* subsp. *zooepidemicus*, an opportunistic pathogen found in many animal species including cattle (Edwards et al., 1988), and *S. suis*, *S. iniae* and *S. canis* typically associated with pigs, fish and dogs respectively (CFSPH, 2005). In addition, humans are reservoir of some *Streptococcus spp.* such as *S. pyogenes* that can be transmitted to animals (reverse zoonosis) causing eventually infection of the udder, contamination of the milk and outbreaks in humans (CFSPH, 2005).

For many streptococcus species found in both humans and animals, the zoonotic potential is uncertain and yet to be elucidated, among these is *S. agalactiae*, a highly contagious obligate parasite of the mammary gland and a major cause of bovine mastitis (Martinez et al., 2000). While the vast majority of human streptococcal infections are attributed to this pathogen, *S. agalactiae* strains recovered from human and bovine sources usually belong to completely separate genetic lineages (Botelho et al., 2018).

Milk and dairy products had a major role in the past; outbreaks of septic sort throat and scarlet fever were numerous prior to the introduction of milk pasteurisation (Macgregor, 1930; Scamman, 1929). Since the ’80, evidence of streptococcal outbreaks (mainly due to *S. zooepidemicus*) involving dairy products have been occasionally reported (Bordes-Benítez et al., 2006; Edwards et al., 1988); most of the recent outbreaks have involved *S. pyogenes* and foods such as salads or eggs, with the source of infection being an infected food handler (FDA, 2012; Greig et al., 2007).

### *Toxoplasma gondii*

| Shed in milk | Yes |
| --- | --- |
| Milk contaminant | No |
| Main source of indirect contamination |  |
| Survive pasteurization | No |
| Severity of illness | Serious |
| Obligate intracellular coccidian protozoan. |  |

*Toxoplasma gondii* is an intracellular parasite causative agent of Toxoplasmosis, one of the most common parasitic zoonoses world-wide (Tenter et al., 2000). Infection in humans is largely subclinical or associated with self-limited symptoms, but can be severe for immunocompromised persons and cause of serious health problems if the parasites are transmitted to the fetus (i.e., congenital toxoplasmosis) in pregnant woman with severe sequelae for the infant (Jones et al., 2003).

Because of the life-cycle of the parasite (Dubey, 2014), infection can be acquired by either ingestion of meat and meat products infected with live *T. gondii* tissue cysts or environmental exposure to sporulated oocysts shed by felids. Although for only about half of the cases it is typically possible to identify the source of infection by means of case–control studies or interviews, the main route of infection seems to be foodborne (Petersen et al., 2010), and mainly due to ingestion of raw or undercooked meat and unwashed fruit and vegetables (Hill & Dubey, 2018; Hussain et al., 2017). The infectious dose in humans is unknown but supposed to be low, experimental data from pigs suggest that as few as 10 sporulated oocyst are deemed sufficient for infection (Dubey, 1986).

The parasite has been detected in camel and donkey milk (Dehkordi et al., 2013; Mancianti et al., 2014), experimentally, it has also been demonstrated that *T. gondii* can be intermittently excreted in goat’s milk and can survive in fresh cheese made by cold-enzyme treatment (Dubey et al., 2014).

While consumption of raw goat milk is often identified as a risk factor for human toxoplasmosis in epidemiological association studies, the risk of acquiring infection by drinking cow's milk, if any, seems to be minimal (Boughattas, 2017).

### *Yersinia enterocolitica*

| Shed in milk | No |
| --- | --- |
| Milk contaminant | Yes |
| Main source of indirect contamination | Environment |
| Survive pasteurization | No |
| Severity of illness | Serious |
| T°C range (growth) | -5-44 °C |
| T°C optimum (growth) | 22-28 °C |
| pH range (growth) | 4.6-10 |
| pH Optimum (Range) | 7-8 |
| Minimum a_w_ (for growth) | 0.96 |

*Y. enterocolitica* is a ubiquitous microorganism; frequently found in soil, water and animals, the pathogen can grow in a variety of foods. In particular, the bacteria is normally present in pigs (Drummond et al., 2012), and pigs are in fact identified as the primary source of human yersiniosis ((Virtanen et al., 2012) and references therein). Typical yersiniosis symptoms include fever, abdominal pain, and diarrhoea (often bloody in young children), fever and appendicitis-like pains are more common in older children and adults. Not life threating, symptoms usually last from 1 to 3 weeks or longer (CDC, 2016).

*Y. enterocolitica* is destroyed by pasteurisation but can growth at refrigeration temperatures, its presence in heat-treated milk products is in fact mainly due to environmental contamination after heat treatments.

The infectious dose is uncertain but likely to be high (i.e. 10^6^-10^8^/g (Fleming & Hunt, 2006)), in the absence of competing microflora, the pathogen multiply rapidly while presence of starter culture or competing microorganisms has an inhibitory effect on the growth of the pathogen.

The presence of presumptive enteropathogenic *Y. enterocolitica* in raw milk, pasteurized dairy products, and asymptomatic handlers has been reported in the US, France, Australia, Brazil, Finland, and China, among other countries.
